# Design for replicability in open-source distributed assistive technology for low-resource settings: a case study of two-piece 3D-printed forearm crutches

**DOI:** 10.64898/2026.02.13.26345756

**Authors:** Alessia Romani, Rebecca Kaaya Nansubuga, Maryam Mottaghi, Danielle Munang, Emily Bow Pearce, Pooja Viswanathan, Thomas Jenkyn, Tarek Loubani, Jacob Reeves, Joshua M. Pearce

## Abstract

**Purpose:** Distributed manufacturing of open-source assistive technology shows potential to offer accessible, affordable, and customizable solutions for users in low-resource contexts. Their real-world adoption, however, depends not only on the availability of openly shared designs but also on their replicability when fabricated in different local contexts. This work investigates the replicability of open-source hardware in assistive technology through a practical design-driven approach, using the development and experimental evaluation of a two-piece open-source forearm crutch as a case study.

**Materials and Methods:** Replicability was considered from early-stage design and evaluated by introducing controlled variations from distributed manufacturing contexts, e.g., material feedstock, manufacturing equipment, and fabrication strategies. Four batches of crutches were fabricated and assembled using virgin and recycled filaments on small-and large-format 3D printers. After a qualitative evaluation, mechanical static load testing was performed following ISO 11334:2007, together with economic analysis.

**Results:** Comparable mean load-bearing and consistent failure behavior were achieved across batches, making them suitable for use in pairs as assistive mobility devices. Limited cost variability was achieved, supporting repairability and product lifecycle extension, while enabling affordable and easy fabrication in different local contexts.

**Conclusions:** Beyond the specific case study, the replicability of open-source hardware for assistive devices needs to be considered as an early-stage design constraint by developing products that allow for variability from local contexts and by including product-specific approaches to assess replicability during development. This design shift can support accessibility and real-world adoption of open-source assistive technology in low-resource settings.

## Introduction

Access to assistive devices in low-resource settings is still challenging due to high costs, limited availability, and reliance on centralized manufacturing (World Health Organization and United Nations Children’s Fund 2022). Distributed manufacturing and open-source hardware (OSH) have been increasingly proposed to improve access to essential products, including assistive products, by enabling local, flexible, low-cost, and distributed production (Bonvoisin et al. 2017; Okwudire and Madhyastha 2021). Physical products based on these principles can be fabricated using widely available tools and technologies, such as low-cost desktop-size 3D printers, by making the source documentation publicly available (Wittbrodt et al. 2013). Their increasing adoption introduces a paradigm shift in engineering design, bringing product design and development into the public domain (Bonvoisin et al. 2017) and opening the design process to a broader audience (Boisseau et al. 2018). Physical products made as OSH aim to increase affordability and accessibility by lowering economic barriers (Pearce 2017), supporting local communities and users (Gwamuri et al. 2014) to customize their products to specific contexts and needs (Boisseau et al. 2018).

This shift led to framing the concept of replicability in OSH, which refers to the ability of an independent user, or prosumer, to make a functional version of a specific OSH within their local context (Antoniou et al. 2021). Contrary to open-source (OS) software, OSH transfers the freedom to study, modify, and distribute products into the physical world (OSHWA 2025), adding additional technical and design challenges to their adoption. For instance, sharing a bill of materials (BOM) and assembly instructions may not be enough to ensure the full reproducibility of an OS physical product (Antoniou et al. 2021). From the literature, replicability emerges as a design challenge that goes beyond the quality of shared documentation, requiring appropriate design choices to simplify the fabrication process and a deeper consideration of the specific context of replication, e.g., available materials, resources, user knowledge, or local makers’ skills (Omer et al. 2024; Antoniou et al. 2022).

To this end, distributed manufacturing further increases the replicability challenge in OSH. Although it enables on-demand and local fabrication of accessible and affordable products, distributed manufacturing also introduces additional risks connected to context-specific variability that are usually not faced in centralized global supply chains (Rayna et al. 2023). OSH products, however, are rarely fabricated under identical conditions, especially across different local contexts. This aspect affects the successful adoption of OSH, even using the same design source and documentation, by introducing intrinsic variability into replication (Antoniou et al. 2021; Gwamuri et al. 2014). As a result, the replicability of OSH is still underexplored in distributed manufacturing contexts. For example, OSH products can be optimized for specific geographical or socio-economic contexts but are hardly replicable in other settings, e.g., when built in low-resource settings (Omer et al. 2024). In these cases, OSH fail to meet practical replicability because constraints from the local context are not properly considered (Omer et al. 2024; Antoniou et al. 2022), for instance, manufacturing limitations, different availability of off-the-shelf components, or heterogeneity in material feedstocks.

Replication challenges introduced by OSH and distributed manufacturing become particularly relevant within specific application sectors, such as assistive technologies and medical devices. Real-world adoption in this context remains challenging, especially in low-resource settings, as functionality, structural performance, and safety are essential requirements for both proprietary and OSH assistive products (Pearce 2020; Stirling and Bowman 2020). Moreover, the need for accessible and affordable assistive and medical devices, such as mobility aids, has become increasingly urgent since global populations are aging. Over 10% of adults worldwide experience mobility-related disabilities (Iezzoni 2003; CDC 2024), and this number is expected to grow (Iezzoni et al. 2001). Chronic or temporary conditions, e.g., back pain, arthritis, and injuries from accidents, are driving a growing demand for adaptive devices, particularly among seniors (Iezzoni et al. 2001). Despite the increasing need, commercial options are often expensive or less adaptable to individual users, limiting accessibility in low-resource settings or different socio-economic contexts (Cubanski et al. 2018; Yanatma 2024; United Nations 2021). OSH has represented a promising solution for affordable medical devices, achieving up to 94% in economic savings compared to commercial options (Ariza and Pearce 2022). Nevertheless, the real adoption of OS assistive devices is strongly linked to the safety and structural reliability of the replicated product, requiring rigorous mechanical and safety standards for their safe use. This shift toward OSH in assistive technologies also lies in the increasing accessibility of distributed digital manufacturing, such as 3D printing (Gershenfeld 2012; Rundle 2014), which has become a powerful medium for fabricating customized, cost-effective goods at a local scale (So et al. 2023; Anderson and Sherman 2007). As a result, an increasing number of essential products, including mobility devices and assistive devices, are locally replicated in libraries, makerspaces, and community fablabs (Moorefield-Lang 2015; Beltagui et al. 2021; Taoheed et al. 2025).

OS approaches to products have further accelerated this transformation (Oberloier and Pearce 2018; Beltagui et al. 2021), reducing costs and improving accessibility (Petersen and Pearce 2017). From the literature, distributed OSH has demonstrated performance comparable to commercial products at a fraction of their cost (Mottaghi et al. 2025; Ariza and Pearce 2022), while allowing for customization and adaptation to specific user needs using available materials and 3D-printed parts (So et al. 2023). Despite these advantages, many of these products implicitly define specific replication conditions (Omer et al. 2024), such as the use of less accessible 3D printers or materials and components available only in specific contexts, which may be difficult to find elsewhere. Replicability, therefore, is often assumed without properly designing or evaluating it, also due to the broad range and heterogeneity of potential local contexts. A critical gap, therefore, remains in ensuring the replicability of OSH within distributed manufacturing contexts, including assistive technology, as replicability is rarely systematically considered and assessed during the design process. At the moment, few studies have analyzed the replicability challenges in OSH (Omer et al. 2024; Antoniou et al. 2021), highlighting the need to define practical and experimental approaches to consider replicability during the development of OSH. Moreover, no previous studies have explicitly focused on the replicability of OS assistive technology, despite the increasing interest in its adoption.

To address these challenges, this study investigates replicability in OSH assistive products through a practical approach, i.e., the design and development process of a two-piece OS forearm crutch as a representative case study. An experimental replicability framework was defined to explore how variations in the distributed manufacturing context influence the replication of the same OS assistive device. Four different batches of crutches were fabricated and assembled, starting from the same design files, developed to be easily fabricated by non-expert users, and introducing variables such as available material feedstock, 3D printing scales, and fabrication strategies. Replicability was assessed through qualitative evaluation of the 3D-printed components and assembled products, mechanical static load testing according to the ISO 11334:2007 standard (ISO 2022), and economic analysis of fabrication costs. The work follows a design-driven approach to improve the replicability of OS assistive products in distributed manufacturing contexts by evaluating patterns of controlled variability and highlighting the key factors affecting the replicability of the specific OSH. The results are presented and discussed in the context of the design process of OS products and assistive technology, leading to improvements that consider their whole product lifecycle, such as disassembly, repairability, repurposing, and lifecycle extension.

## Materials and methods

Figure 1 shows the experimental workflow of this work. An OS 3D-printable forearm crutch was developed as a case study to assess the replicability and repairability of OS assistive products in distributed manufacturing and low-resource contexts, as explained in the next sub-section. After the first design iteration and preliminary prototypes of the product were made, the 3D model was further refined to optimize the fabrication of the 3D-printable components, their assembly, and the overall repairability of the crutches. An experimental framework for the replicability assessment was then defined by organizing fabrication, testing, and analysis into different batches and testing conditions, aiming to simulate a representative distributed manufacturing context for the developed product. The different 3D printing components and off-the-shelf parts were fabricated and assembled to prepare the batches, which reflect variations in using different material feedstock sources and manufacturing setups. After a visual and qualitative comparison, the assembled products were then tested to evaluate the mechanical properties under static loading conditions, using the ISO standard (ISO 2022) as a reference loading condition for consistent comparison across batches and to assess variability in replication in distributed manufacturing contexts. The replication costs were then assessed through an economic analysis of the different batches, comparing variations in material and manufacturing costs.

**Figure 1.**
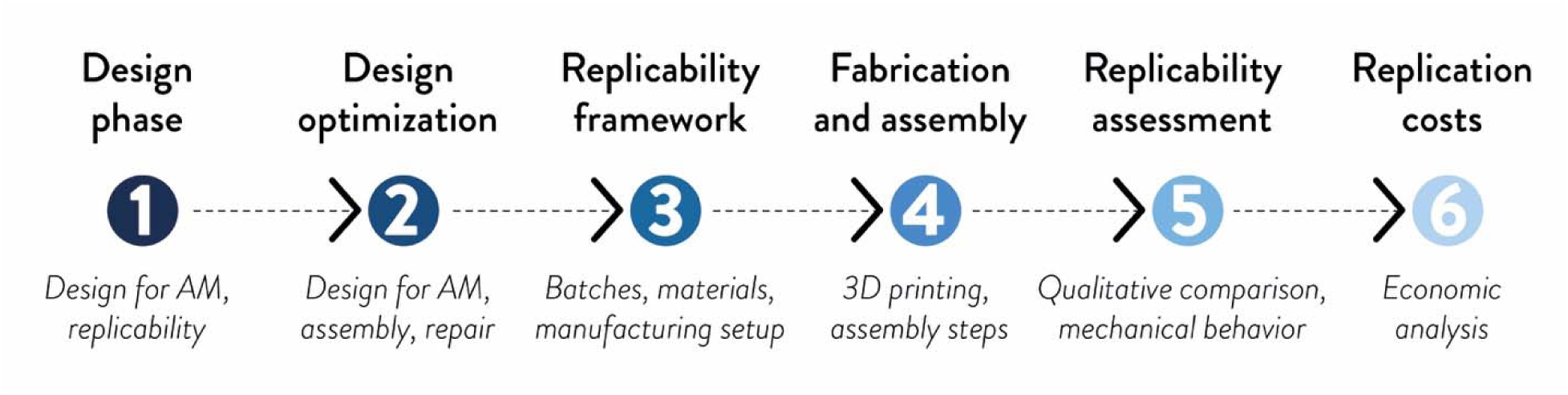
Experimental workflow followed for the replicability assessment of the OS forearm crutches in distributed manufacturing contexts: (1) design phase according to the design for additive manufacturing (DfAM) principles and selected context; (2) design optimization for AM, assembly, and repair; (3) definition of the experimental replicability framework; (4) fabrication and assembly of the different crutch batches; (5) replicability assessment through qualitative comparison and mechanical tests under static loading; and (6) assessment of the replication costs.

### Design of the forearm crutch for low-resource distributed manufacturing contexts

This work focuses on the replicability of OS assistive products in low-resource distributed manufacturing contexts, such as humanitarian settings or remote areas, to support their adoption in real-world scenarios. These contexts require accessible products following the principles of open design and distributed manufacturing, e.g., in the healthcare and medical sector, that meet adequate quality in terms of functionality, safety, and feasible replication in distributed local contexts (Ariza and Pearce 2022). Among those, forearm crutches are a widely used class of assistive and medical devices following specific safety and ergonomic requirements, representing a relevant example of potential OSH to be replicated, assembled, and used in different real-world contexts, e.g., geographical or socio-economic contexts. For these reasons, they are suitable to assess practical aspects behind the development and replication of OS assistive products for their real adoption through distributed manufacturing.

The design of the OS crutch, a two-piece forearm type, is a redesign of a previous OSH that could be manufactured in one component but could not be manufactured on small-scale 3D printers (Mottaghi et al. 2025). This new version was guided by a set of constraints related to these contexts, mainly limited availability of manufacturing equipment or materials, or variations in available off-the-shelf components, e.g., different standard unit systems or local supply chains. In detail, the custom components were designed according to the principles of Design for Additive Manufacturing (DfAM) (Pradel et al. 2018; Borgianni et al. 2022), aiming to: (i) fit within the build volume of low-cost desktop-size fused filament fabrication (FFF) 3D printers, i.e. 200×200×200 mm; (ii) have custom parts replicable using widely accessible material feedstocks for 3D printing, according to local availability; (iii) include features that help the user to manufacture or replicate the parts regardless of the expertise level of who is replicating them; (iv) use simple and accessible off-the-shelf components, when standard components are needed; (v) maximize the possibility of adapting the product without using CAD software; (vi) limit the need for post-processing of the custom parts; and (vii) enabling repair and repurpose of the product by easily replacing or changing specific components.

The 3D model of the crutch was developed using Onshape 1.157 (Cambridge, MA, US) (Onshape 2025). The 3D-printable parts were designed to be easily fabricated regardless of the expertise level of those in charge of the replication by: (i) minimizing geometry complexity; (ii) avoiding the use of supports; (iii) including large and stable surfaces as contact areas with the build plate; and (iv) using overhangs achievable with common desktop-size FFF 3D printers and standard parameters, e.g., nozzle sizes and layer heights.

The assembly of the two-piece forearm crutch is visible in Figure 2a, and it consists of three main sub-assemblies, i.e., the cuff, the handle, and the body. It uses both custom and off-the-shelf parts (Figure 2b), with 3D-printed parts fabricated from either virgin filament or recycled plastic, depending on material availability, resource limitations, and cost considerations specific to the replication context, e.g., local availability of off-the-shelf parts or variation in standards.

**Figure 2.**
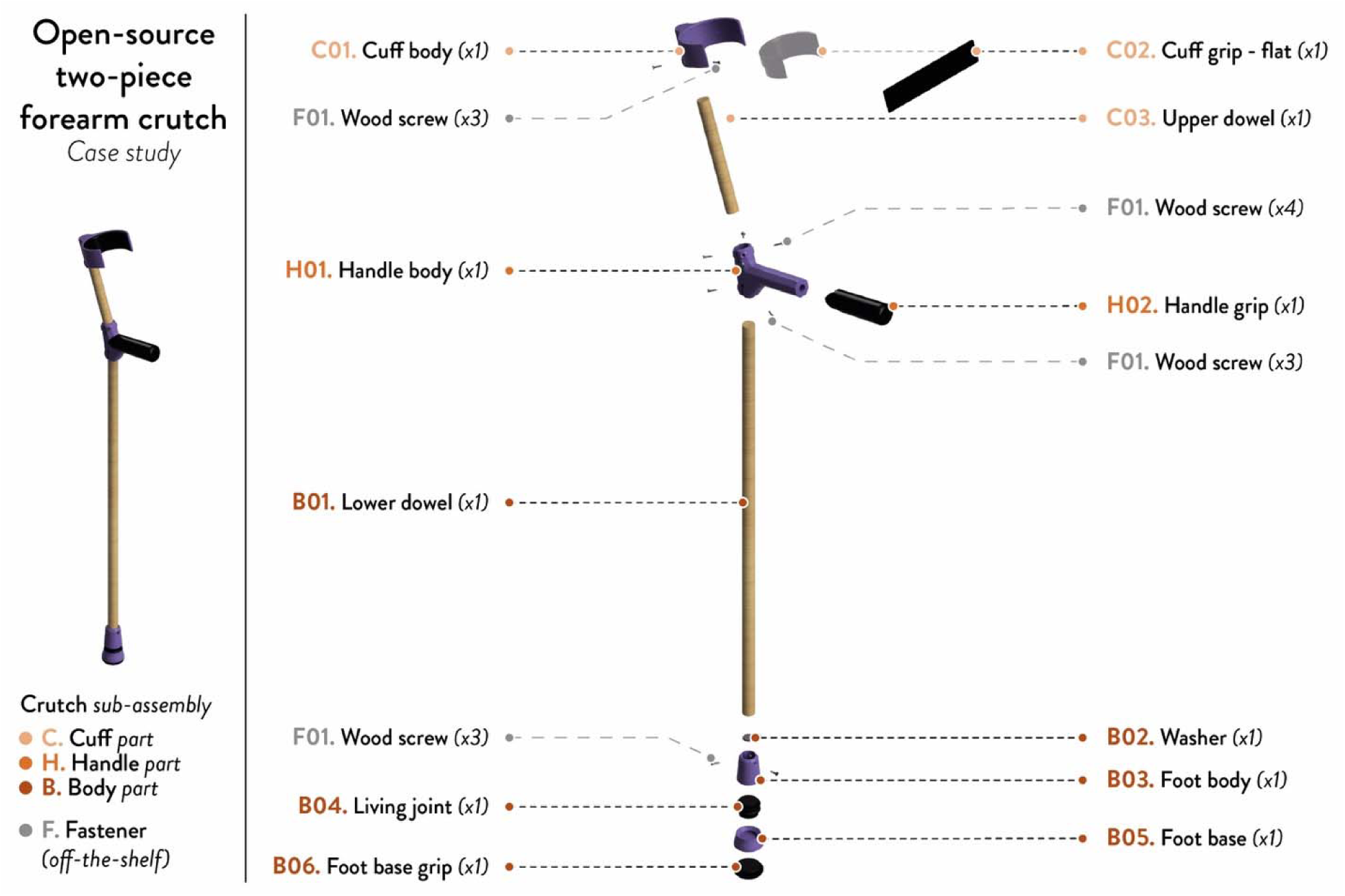
OS two-piece forearm crutch: overall assembly with the sub-assemblies (on the left) and exploded view of the assembly with the different components and quantities (on the right).

Focusing on the individual parts, the handle (H01) and cuff (C01) were designed to ensure comfort and maintain proper wrist and hand alignment, according to the ISO 11334-1:2007 standard (ISO 2022). Anthropometric dimensions of the forearm cuff, such as internal depth, width or and height, are adjustable to fit individual user needs. The overall height of the crutch and forearm support can be adjusted by measuring and cutting the hardwood dowels (C03 and B01), simplifying customization without the use of digital tools. Grips for the handle (H02) and cuff (C02) were designed to improve comfort and enable small adjustments of the dimensions, whereas flat washers (B02) were included between the wooden dowel and 3D printed foot to absorb shocks during use, compensate for minor misalignments, and ensure even load transfer across the joint. The foot components, i.e., foot body (B03), living joint (B04), foot base (B05), and foot base grip (B06), include a combination of rigid and flexible 3D-printed components to ensure stability and improve ground contact. Parts were also developed considering either easy disassembly for reuse, repair, repurpose, or quick adaptations, e.g., for other users. In some cases, additional 3D models are provided as alternative parts to facilitate their disassembly, such as for the foot components (Figure S7 and Figure S8, Supplementary Information). The BOM is available in the Supplementary Information (Table A1, Section S1). The OSF repository (Romani et al. 2026) includes 3D models of the assembly and custom parts (STEP format), as well as STL files of the 3D-printable parts.

### Experimental replicability for distributed manufacturing contexts

The replicability of the OS forearm crutch, selected as a case study, was analyzed by defining a set of sample batches that represent plausible variations in the distributed manufacturing of the parts. This approach aimed to assess if the same OS design can be successfully replicated and used when some of the key aspects of the manufacturing process change according to the specific local contexts, such as material feedstock origin, 3D printer scale, and production settings. Different batches of samples were therefore defined to reflect realistic replication scenarios, including: (i) the use of virgin or recycled filament feedstocks; (ii) small- or large-format FFF 3D printers; (iii) the fabrication of single or simultaneous parts; and (iv) the production of the same product at different fabrication times.

Table 1 shows the different batches defined for the replicability assessment. In detail:

- **Batch B01** (SF_VPETG) is the set of crutches fabricated using virgin polyethylene terephthalate glycol (PETG) filaments for rigid parts and thermoplastic polyurethane (TPU) filaments for flexible components on a desktop-size FFF 3D printer, fabricating each component individually. This batch was initially made of five samples (n=5).
- **Batch B01b** (SF_VPETG) is the replication of the same conditions of B01 at a different time, extending the overall amount of the batch to ten (n=10) by adding these last five samples. This second fabrication batch creates a direct comparison between smaller and broader replications of the same product, evaluating the variability of the replication under unchanged fabrication conditions.
- **Batch B02** (SF_RPETG) uses recycled commercial PETG feedstock, keeping the same TPU material, small-format 3D printing apparatus, and single-part fabrication strategy. This batch (n=5) simulates a distributed manufacturing context where recycled filaments are preferred due to costs, availability, or circularity considerations.
- **Batch B03** (LF_VPETG) switches the replication to large-format FFF 3D printers, which can be available in local collaborative manufacturing contexts, e.g., fab labs and makerspaces. In this case, the samples (n=5) are manufactured using the same virgin PETG materials of B01, and simultaneous fabrication of the parts was selected to fabricate the components due to the area of the build plate, simulating a possible scaling-up pattern of these products.

**Table 1.** Batches of the crutch samples for the experimental work (ID, number of samples per batch, and description), together with the main 3D printing apparatus and material feedstocks used for the replicability assessment in distributed manufacturing contexts.

| Batch N. and ID | N. | Batch description | 3D printing system | Rigid 3D printed parts | Flexible 3D printed parts |
| --- | --- | --- | --- | --- | --- |
| B01. SF_VPETG (n=5) | 5 | 3D printed parts (virgin filaments) from desktop-size 3D printers, printed one at a time (original batch) | Small-format FFF | Virgin PETG | Virgin TPU 90 shore A |
| B01b. SF_VPETG (n=10) | 5+5 | 3D printed parts (virgin filaments) from desktop-size 3D printers, printed one at a time (with five additional specimens fabricated at a different time) | Small-format FFF | Virgin PETG | Virgin TPU 90 shore A |
| B02. SF_RPETG | 5 | 3D printed parts (recycled filaments) from desktop-size 3D printers, printed one at a time | Small-format FFF | Recycled PETG | Virgin TPU 90 shore A |
| B03. LF_VPETG | 5 | 3D printed parts (virgin filaments) from large-format 3D printers, printed simultaneously | Large-format FFF (Modix 3D Printer 2022) | Virgin PETG | Virgin TPU 90 shore A |
| B04. RF_RPETG | 5 | 3D printed parts (recycled filaments) from large-format 3D printers, printed simultaneously | Large-format FFF | Recycled PETG | Virgin TPU 90 shore A |

- **Batch B04** (LF_RPETG) combines the use of a large-format FFF apparatus and simultaneous fabrication of multiple parts with the use of recycled PETG, reflecting the possible use of recycled filaments at a larger scale on this set of samples (n=5).

The same design files, assembly procedure, and off-the-shelf components were used to fabricate all these sample crutches, ensuring that the observed results could be linked to replication variables. The same small- and large-format 3D printers were used to fabricate the parts, as well as the same virgin and recycled PETG filaments. Only virgin TPU was used for the soft parts, which were fabricated only on the small-format 3D printer due to the challenges of recycling flexible filaments and printing TPU on large-format 3D printers. As explained in the next sub-sections, replicability was evaluated through: (i) a qualitative assessment of the 3D printed parts and assembled products; (ii) a mechanical test of the crutches under static load conditions; and (iii) an economic analysis of the different replicas.

### Fabrication of the 3D printed components

According to Figure 2 and the BOM (Table S1, Supplementary Information), the custom components of the OS crutch can be fabricated using a desktop-size FFF 3D printer, which represents a widely accessible fabrication tool in distributed manufacturing contexts. For this study, the 3D-printed parts were fabricated with both a small- and a large-format 3D printer, enabling the replication assessment across different apparatus scales, i.e., Prusa i3 MK3S (Prusa Research, Prague, Czech Republic) (Prusa Research 2025) and Modix BIG-Meter (Modix GmbH, Koblenz, Germany) (Modix 3D Printer 2022). Both 3D printers were equipped with a 0.8 mm nozzle, and the gcode files were prepared with the OS slicing software PrusaSlicer. As shown in Figure 3, the building strategy of the components changed with the scale of the 3D printers, simulating real-world conditions for manufacturing these parts with both 3D printers, e.g., one part at a time (Figures 3a and 3b) or simultaneous fabrication of multiple components (Figures 3c and 3d).

**Figure 3.**
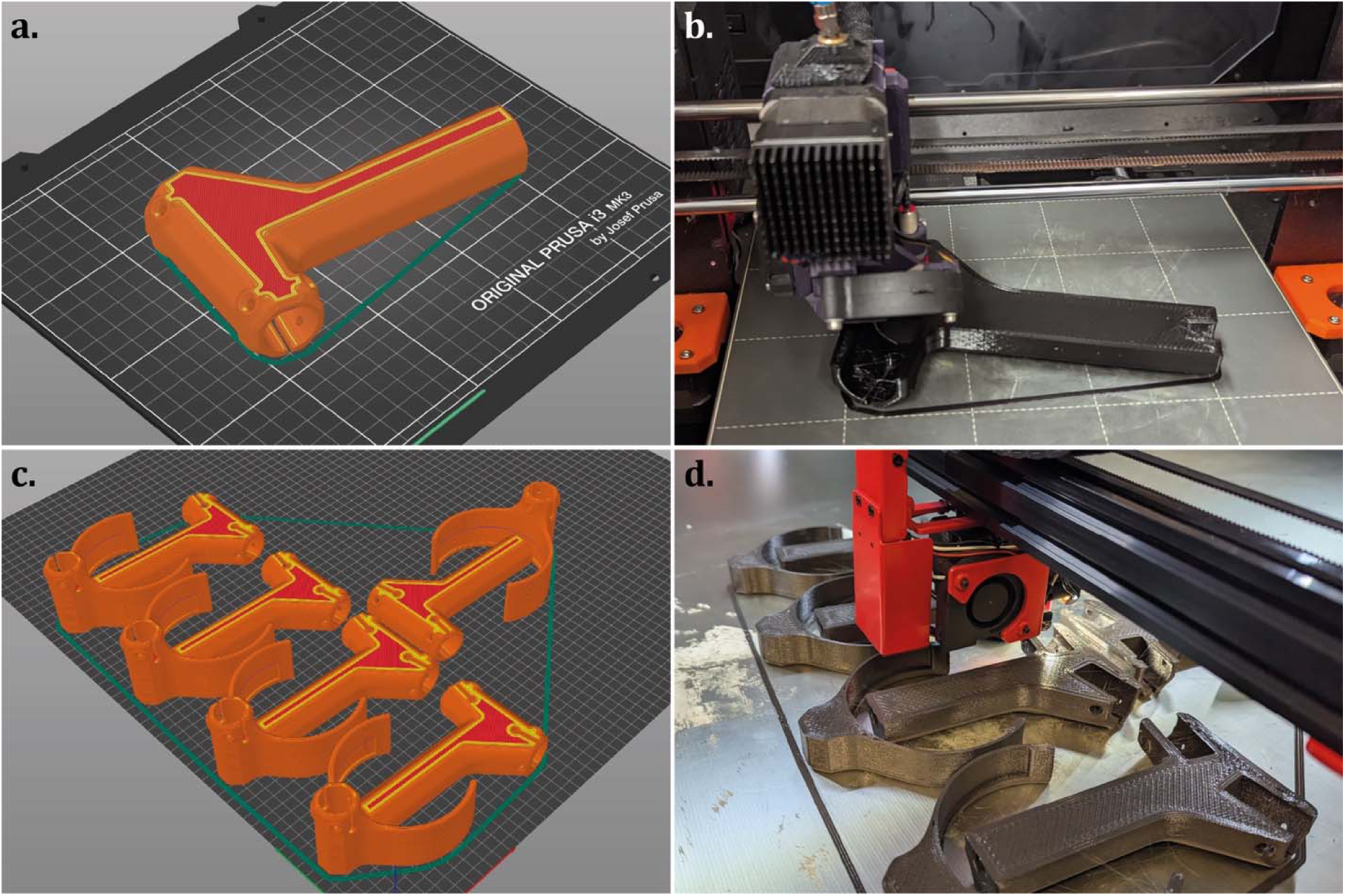
Building strategy of the 3D printed components according to the 3D printer scale: (a) orientation of the handle for small-scale 3D printers (single component) and (b) fabrication of the corresponding gcode; (c) orientation of multiple handles for large-scale 3D printers (simultaneous fabrication) and (d) fabrication of the corresponding gcode.

Table 2 shows the main 3D printing settings used for each sample batch, according to the selected 3D printer and material. The rigid 3D-printed parts were fabricated using either virgin PETG (ELEGOO US 2026) or recycled PETG (Spectrum Filaments 2022), depending on the specific batch. Fully dense parts were fabricated from PETG to ensure high mechanical strength, strong layer adhesion, and good surface quality. Increased infill overlap and flow rate were selected to reduce interlayer voids, which can cause premature failure and localized cracking. Virgin TPU 90 Shore A filaments were used for all the flexible parts (PolyMaker 2026), and different levels of flexibility were designed by varying the infill percentage, using a higher-hardness filament (90 Shore A), which can be easily 3D printed by entry-level users.

**Table 2.** 3D printing parameters for the 3D printed parts of the OS crutch, divided according to the 3D printer formats, materials, and specific batch.

| Parameters | Unit | Prusa MK3S<br>(Small-format FFF) |  |  | Modix Big meter<br>(Large-format FFF) |  |
| --- | --- | --- | --- | --- | --- | --- |
| Material | // | Virgin PETG | Recycled PETG | Virgin TPU 90 Shore A | Virgin PETG | Recycled PETG |
| Parts | // | C01, H01, B03, B05 |  | C02, H02, B02, B04, B06 | C01, H01, B03, B05 |  |
| Batch | // | B01, B01b | B02 | All | B03 | B04 |
| Nozzle diameter | mm | 0.8 | 0.8 | 0.4 | 0.8 | 0.8 |
| Extruder temperature | °C | 230 | 230 | 230 | 230 | 230 |
| Bed temperature | °C | 80 | 80 | 50 | 80 | 80 |
| Speed | mm/s | 40-60 | 40-60 | 20 | 40-60 | 40-60 |
| Infill (percentage) | % | 100 | 100 | 10 (100 for part B02) | 100 | 100 |
| Infill (overlap) | % | 35 | 35 | 10 | 35 | 35 |
| Flow (percentage) | % | 105 | 105 | 115 | 105 | 105 |
| Layer height | mm | 0.3 | 0.3 | 0.3 | 0.3 | 0.3 |
| Perimeters | // | 3 | 3 | 2 (1 for part H02) | 3 | 3 |
| Top/bottom layers | // | 3 | 3 | 4, 3 | 3 | 3 |

Furthermore, the flexible parts were designed in a flat configuration, e.g., cuff grip, or alternative flat geometries were provided to simplify fabrication or customization, e.g., handle grip. The gcode and 3mf files of the 3D printed parts, organized by batch, are available in the OSF repository (Romani et al. 2026), allowing for tuning comparable fabrication settings.

### Selection of components and assembly of the crutches

The other components were selected according to their suitability for low-resource distributed manufacturing contexts, easy replacement, and availability. Solid cylindrical hardwood dowels with a 22.3□mm (7/8”) diameter were used for the structural framework, i.e., the upper and lower dowels (C03 and B01). Hardwood was chosen for its lower environmental impact compared to aluminum, which is commonly used in commercial forearm crutches. Compared with aluminum alloys, the carbon footprint of wood is approximately one-fourth that of aluminum in similar structural applications, such as window frames (Sinha and Kutnar 2012). The substantial carbon emissions associated with primary aluminum production, averaging around 16 tons of CO□per ton of aluminum in 2022 (Carbon Chain 2022), further support the environmental benefits of choosing wood. Considering the selected context, hardwood is also readily available at hardware stores or can be replaced with alternative tools, e.g., canes or broomsticks, facilitating repair and local substitution of components. At their end-of-life, the dowels can be composted or reused, reducing environmental impacts. Recycled plastics also represent a promising alternative, offering up to 79% reductions in energy consumption and 67% reductions in greenhouse gas emissions compared to virgin plastics, depending on polymer type and recycling strategy (Tonini et al. 2021; The Association of Plastic Recyclers 2018). The versatility of hardwood dowels extends to their mechanical performance, which can vary depending on the specific species used. Hardwoods, e.g., basswood, maple, and oak, generally provide superior mechanical strength compared to softwoods, e.g., cedar, pine, or spruce, due to their higher density and cellular structure (Arriaga et al. 2023). In this study, birch hardwood dowels were selected for their widespread availability, dimensional consistency, good strength-to-weight ratio, workability, and compatibility with 3D-printed components. These aspects make it a representative choice for the intended application and the simulated distributed manufacturing context.

To ensure compatibility with users in a high anthropometric percentile (95^th^ adult males) and simulate worse-case loading scenarios (Brauer 2005), the lower dowel (B01) is initially cut to 800 mm, achieving a handgrip-to-floor distance of 880 mm. Additionally, a 210 mm dowel (C03) is cut to connect the handle and cuff sections, creating a gap between the handle and the cuff of 240 mm, meeting the requirements of the 95^th^ male percentile for a correct distance between the elbow and the cuff. Once this maximum configuration is validated, the dowels can be cut to fit individual user needs and anthropometric measurements, e.g., reaching the 5^th^ percentile for adult women or children’s anthropometrics, supporting adaptability.

The assembly only requires wood screws as fasteners, without the need for glue or adhesives. According to the BOM (Table S1, Supplementary Information), #6 x 5/8” flat head wood screws (imperial standard) were selected for their easy availability and replaceability with metric alternatives according to the specific geographical context, without affecting the assembly or functionality, e.g., M4 x 16 mm countersunk wood screws. The different batches of crutches were assembled according to the detailed assembly instructions and reference images available in the Supporting Information, Section S1. Complete assembly 3D models and BOM are also available in the OSF repository (Romani et al. 2026).

### Mechanical static load tests

Mechanical static load tests were performed to demonstrate the replicability of the OS forearm crutch across different distributed manufacturing conditions, according to the batches defined in the previous sub-sections. These tests were also conducted to determine the failure point and the overall performance of the different batches, e.g., for everyday use, to evaluate the variability and consistency in mechanical behavior. The testing protocol was adapted from the static loading method outlined in Section 5.6 of ISO 11334-1:2007 (ISO 2022), which specifies that crutches must support a vertical load of 1000 N ± 2% under the simulated weight of a 100 kg user for 10 seconds without permanent deformation or structural failure.

For testing, a custom OS mechanical test rig was designed and built, using both custom parts and off-the-shelf components, as shown in Figure 4a. This rig is a redesign of a previous version (Mottaghi et al. 2025), modified to allow for easier replication and reduce the need for custom components different from 3D printed parts. The rig aims to replicate realistic biomedical load paths during the use of the tested crutch. It is made of a simulated hand connected to the handle, a wrist joint, a simulated forearm, a 3D-printed elbow joint, and a central jig body to be mounted on the hydraulic press. Its assembly transfers compressive force from the back of the cuff through the forearm and elbow structures to the handgrip. According to Figure 4b, the hand was clamped at the far end of the handle to prevent reinforcement effects and, at the same time, to ensure the crutch remained free to flex and pivot in multiple directions. A swiveling joint beneath the cuff provided multi-directional rotation, i.e., at least 15°, while the forearm-hand hinge allowed for natural forward-backward and minor lateral motion, approximating realistic user kinematics. Details on the design, fabrication, assembly, and use of the testing rig, together with the full BOM, are available in Supplementary Information, Section S2. The OSF repository contains the 3D models and 3D-printing files for the custom parts (Romani et al. 2026).

**Figure 4.**
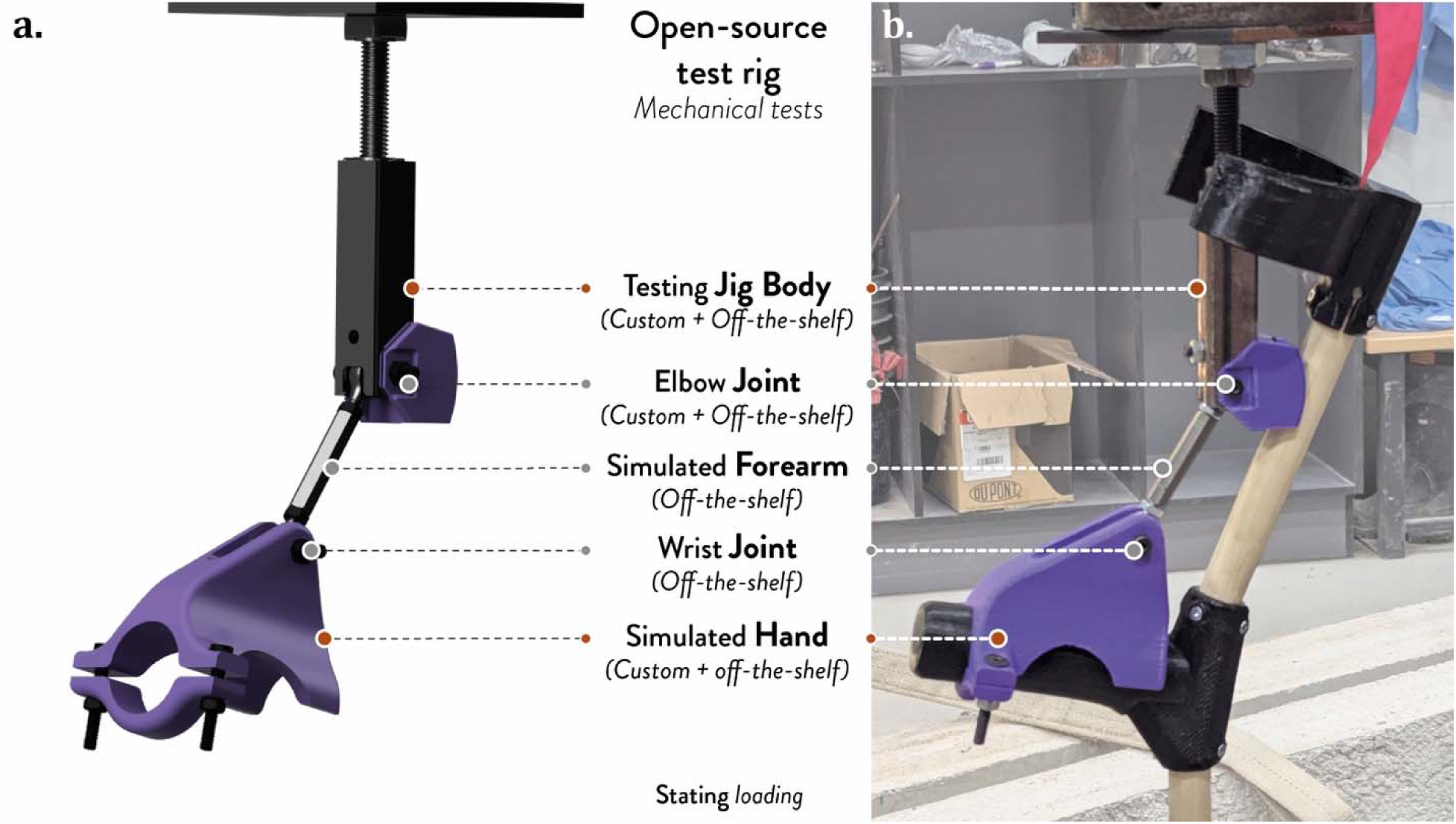
OS custom mechanical test setup (rig) used for the static loading test, adapted from (Mottaghi et al. 2025): (a) 3D model of the assembly and (b) testing setup mounted on the hydraulic press.

Each crutch was secured on the testing rig, which was mounted on an MTS hydraulic actuator with 250□kN capacity and 150□mm stroke that operated in both force-controlled and displacement-controlled modes (MTS System Corporation, Edein Prairie, MN, US) (MTS Systems 2021). The foot base was also aligned to the mounting system before starting the test to ensure proper load transfer during the test, and no slipping occurred between the crutch foot and the test platform, or between the simulated elbow and the rig, to have consistent load application. The actuator applied a vertical compressive load at a rate of 50□mm/min, reaching the 1000□N static load within ∼30s, according to the ISO standard. After holding this load for 10s, loading continued at the same rate until failure. In this work, failure was defined as the first sign of a drop in force with continued displacement, indicating fracture or loss of structural integrity in the dowel or the 3D-printed components.

The compressive force-displacement curves from the experimental tests were plotted to compare the mechanical behavior and variability across the tested batches, after subtracting the initial preload registered by the press during the pretest positioning of the samples. The curves were normalized and linearly interpolated to calculate the mean curves for each batch, which were interrupted after 80% of the samples failed in each batch, allowing comparison between samples with different absolute displacements and failure points. The data were then used to calculate the mean values and standard deviations of the maximum compressive loads and to plot the mean force-displacement curves for each batch. These mean maximum compressive loads were then converted from Newtons to kilograms to allow for direct comparison to bodyweight statistics. Safety factors were defined as the ratio of load to bodyweight using reference adult body masses from Canadian national statistics, i.e., 86.4 kg for men and 72.1 kg for women (Colley et al. 2025). Two loading scenarios were considered for the specific application: single-crutch use, assuming a more conservative context, and use in pairs, assuming equal load distribution between the two crutches, i.e., half of the bodyweight on each device, thereby multiplying the safety factors by two. A qualitative analysis of the fracture points was carried out to identify recurring failure locations and aspects connected to the variability across the batches. The dataset with the experimental data, normalization, interpolation, and mean values is available in the OSF repository (Romani et al. 2026).

### Economic analysis

A cost analysis of the developed OS crutches was performed to assess the affordability and its variability according to the different distributed manufacturing conditions selected for the experimental work. The analysis considered the different material feedstocks and 3D printing scales used for the tests, estimating the fabrication costs for each batch. The analysis considered all the component costs, either custom or off-the-shelf, and excluded labor costs, taxes, and administrative or disposal fees.

The fabrication costs of the 3D-printed custom parts included material and energy costs, calculated from nominal 3D-printing times, weights, and energy consumption. Time and weights were estimated through the gcode files, using the densities of the filaments provided by the manufacturers, i.e., 1.29 g/cm^3^, 1.23 g/cm^3^, and 1.12 g/cm^3^ for virgin PETG (ELEGOO US 2026), recycled PETG (Spectrum Filaments 2022), and virgin TPU Shore A (PolyMaker 2026), respectively. Material costs were 13.86 USD/kg for virgin PETG, 9.71 USD/kg for recycled PETG, and 53.52 USD/kg for TPU. The energy costs were calculated using the power consumption declared by the manufacturers, i.e., 80 W for Prusa MK3S and 1650 W for Modix One (heated bed and 3D printer electronics), the 3D printing times, and the electricity costs, using the data provided by the Ontario Energy Board as a reference for this work, i.e., 0.089 USD/kWh (Ontario Energy Board 2025).

The total cost of the different batches of OS crutches included all the components in the BOM (Table S1, Supplementary Information), including the wooden dowels (7.18 USD per 1219.2 mm or 48’’) and fasteners (0.07 USD/unit), converted in USD for the analysis. The costs for the set of two crutches and the percentage costs for the four batches were also calculated, starting from the total cost. Detailed costs are included in the Supplementary Information, Section S4, and the spreadsheet is available on OSF (Romani et al. 2026).

## Results and Discussion

### Replicability of the OS crutch for distributed manufacturing contexts

The replicability of OS products in low-resource distributed manufacturing contexts was investigated through a new OS two-piece forearm crutch, selected as a case study. By using the same design source, assembly procedures, and off-the-shelf components, the different batches show how specific variations in material feedstock source, 3D printing scale, and fabrication strategy influence the replication of the same OS product in distributed manufacturing contexts at the level of specific parts or overall product assembly. This choice allows evaluating if the same assistive device fabricated in different local contexts can ensure functional consistency and assembly feasibility despite common variations in the fabrication setup.

The fabrication of the rigid PETG parts resulted in geometrically consistent components at the macro level. As shown in Figure 5, the fabrication of the custom components was successfully achieved with both virgin (B01, B03) and recycled filaments (B02, B04), using the same gcode settings on the same 3D printing scale and without the need for geometry modification or changes in the 3D model files. Minor differences were noticed between virgin and recycled PETG at the surface level or in localized overhang or bridging areas, e.g., cuff grip slot or foot base overhang. A similar trend was observed in comparing parts from small- or large-format 3D printing. Parts fabricated simultaneously on a large-format system (B03, B04) show more visible defects, such as stringing or wider extrusion marks, compared to the fabrication of single parts on desktop-size 3D printers (B01, B02). These differences, however, did not result in warping, detachments, or other critical structural issues in the replicated parts, thanks to the design and fabrication choices made during development. This result demonstrates the relevance of considering fabrication constraints from the early stages of the design process, thanks to DfAM (Pradel et al. 2018), such as facilitating replication by non-expert users (Antoniou et al. 2021) through support-free geometries or components with a clearly visible building orientation suggested by the shape, e.g., the wide flat surfaces of the cuff and handle body. It also shows the impact of early-design decisions in facilitating replicability, e.g., mitigating manufacturing variability, reducing the need for geometry modifications, or expert fabrication skills.

**Figure 5.**
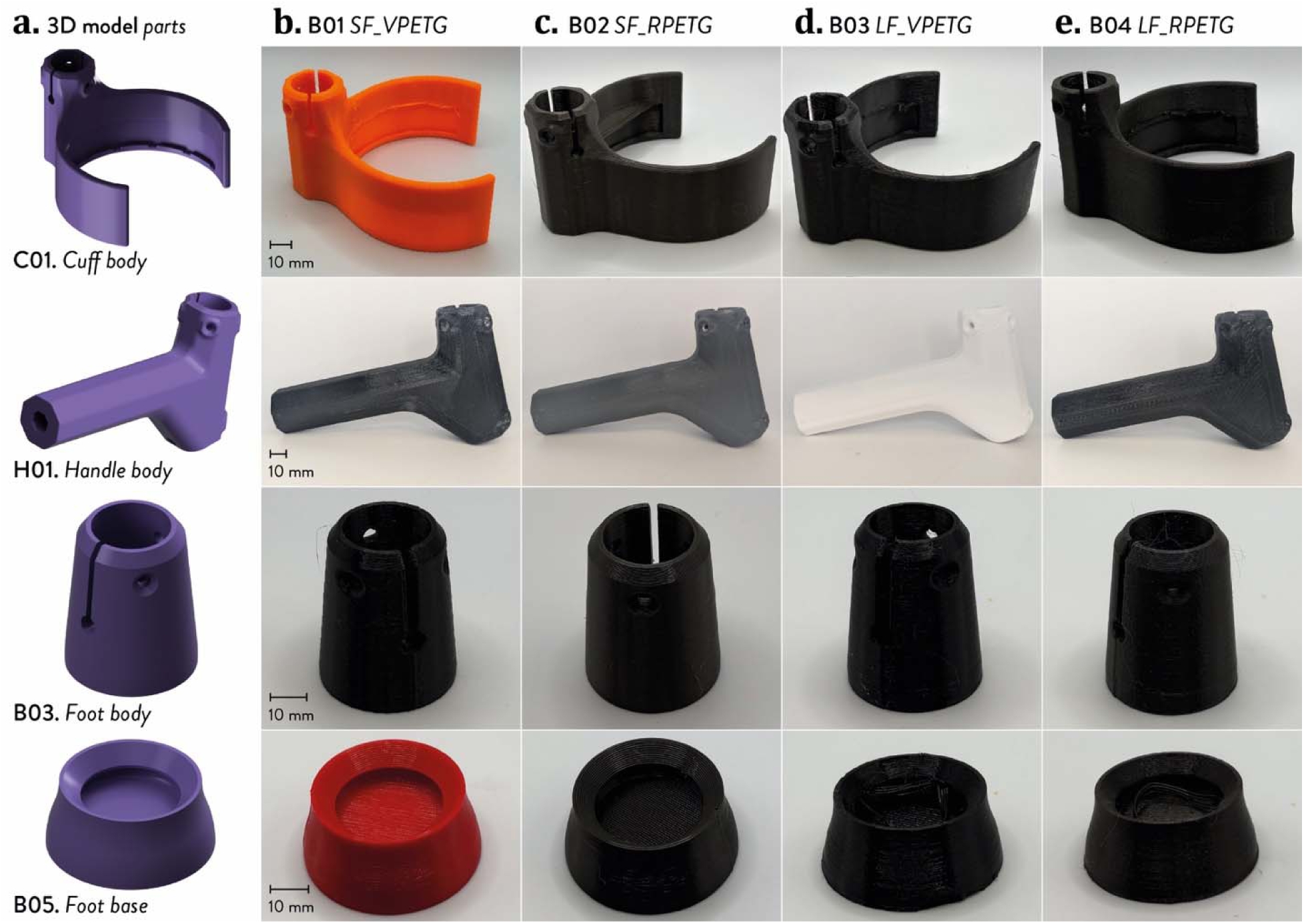
3D printed parts of the crutches made with PETG filaments: (a) 3D models; 3D printed parts of batches (b) B01 (small-format 3D printer and virgin material), (c) B02 (small-format 3D printer and recycled material), (d) B03 (large-format 3D printer and virgin material), and (e) B04 (large-format 3D printer and recycled material).

Flexible TPU components were fabricated only on desktop-size 3D printers. According to Figure 6, designing flat bendable or support-free geometries, combined with the use of semi-flexible TPU filaments with variable infill percentages, resulted in parts with tunable flexibility and, at the same time, easy replication on different systems, e.g., avoiding low Shore A hardness filaments, known to be more complex to be 3D printed for non-expert users, and preferring the customization of the material-infill void ratio. This choice reduces barriers for entry-level users and allows them to use more accessible materials, making replication easier.

**Figure 6.**
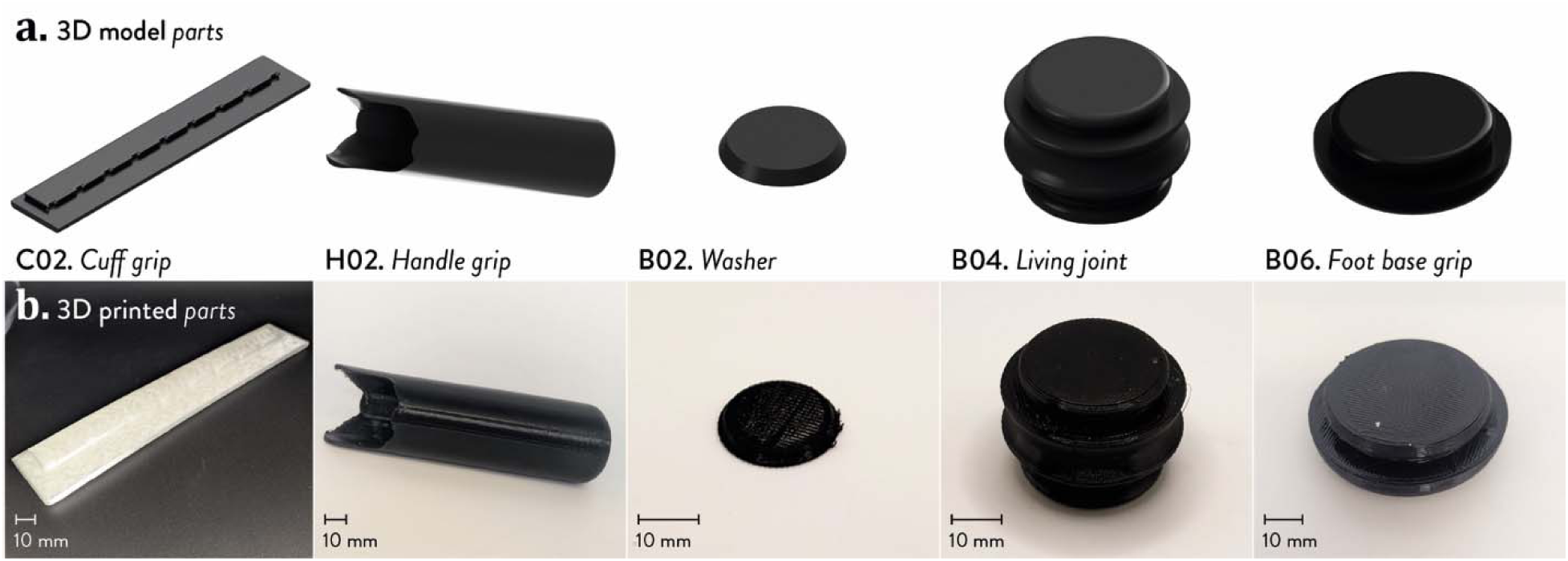
3D printed parts of the crutches made with TPU 90 Shore A filaments: (a) 3D models; and (b) 3D printed parts (small-format 3D printer).

The assembled crutches from the different batches were visually and geometrically comparable (Figure 7). The assembly procedure remained unchanged, without requiring any batch-specific geometrical or alignment adjustments. Maintaining assembly consistency across batches is crucial for these products, where improper alignment or assembly can affect usability and safety. Overall, the simulated distributed manufacturing context demonstrated that the OS crutch can be replicated across different fabrication conditions, keeping assembly feasibility and functional equivalence at the product level during use (Figure 8). Moreover, the repeated part fabrication and assembly for the different batches led to further design refinements, leading to the development of alternative versions of selected 3D-printed components to facilitate disassembly and replacement, e.g., foot rigid parts. This step further supports circular economy practices for distributed OS products (Kane et al. 2018; Romani et al. 2023), such as repair or repurpose, as well as their lifecycle extension, e.g., adapting the dimensions of a second-hand OS crutch to fulfil the needs of next users. These alternative versions of the parts are in the OSF repository (Romani et al. 2026).

**Figure 7.**
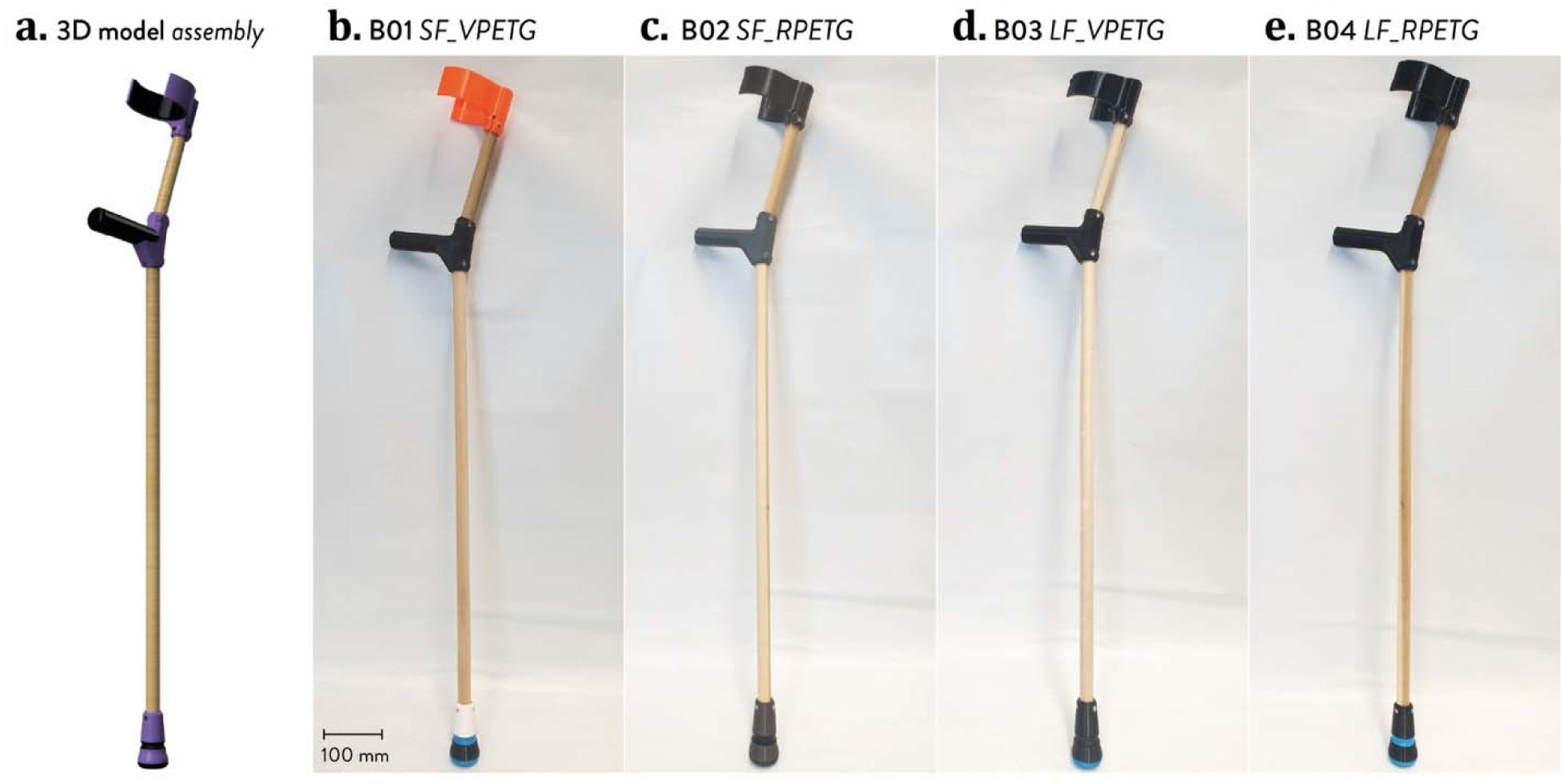
Assembled crutches: (a) 3D model; batches (b) B01, (c) B02, (d) B03, and (e) B04.

**Figure 8.**
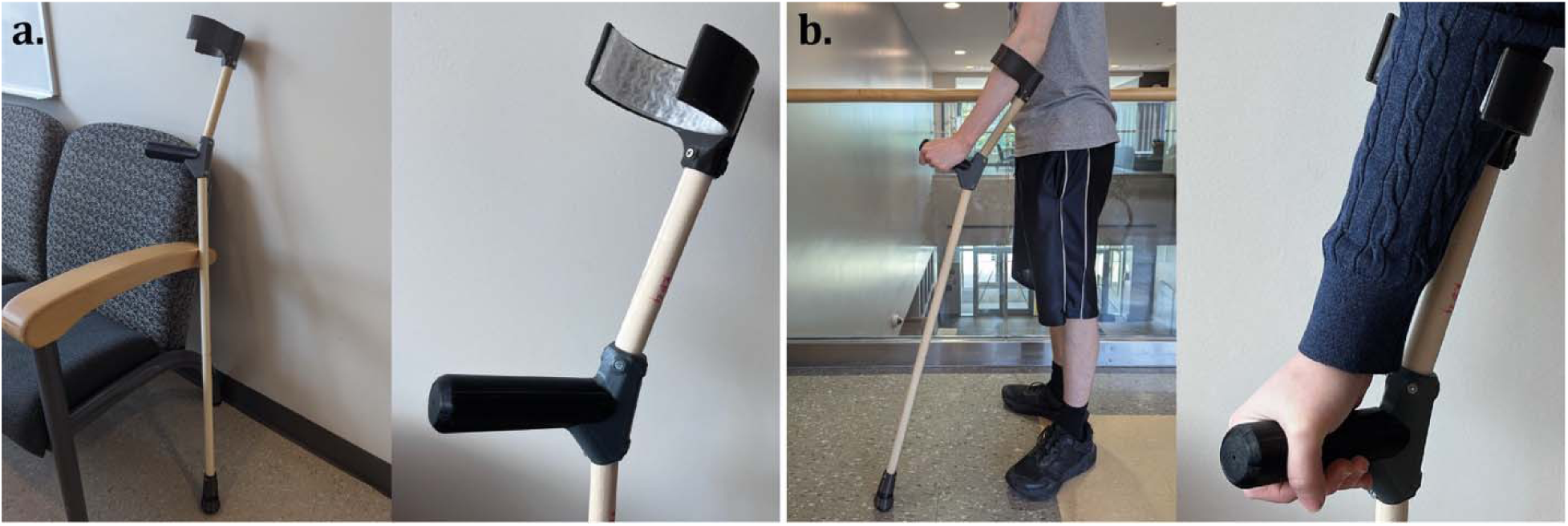
OS 3D printed crutch: (a) assembled product in real context; and (b) product in use.

### Mechanical static load tests

The mechanical tests evaluated the replicability of the OS forearm crutch in different distributed manufacturing settings from a structural and functional perspective. The dataset is available on OSF (Romani et al. 2026).

Figure 9a shows the mean compressive load-displacement curves from the experimental tests for the four different batches of crutches, whereas the corresponding curves are reported in Figure S11 (Supplementary Information, Section S3). In general, the batches exhibited comparable mechanical behavior, with differences in displacement at the load peak, maximum load values, and the range of standard deviations in the mean curves. Significant differences in the mean curve were observed for batch B03, which used 3D-printed components made of virgin PETG on a large-format 3D printer with simultaneous fabrication of multiple components, reaching lower compressive loads but higher displacements before failure. Higher variability in the standard deviations of the mean curves is visible in batches B02 and B04, which represent the sets made with recycled 3D printing material feedstocks. Although the mechanical properties of the crutches are also influenced by other components, e.g., wooden dowels, and the overall assembly quality, this increased variability is usually expected in recycled thermoplastics, which are affected by the recycling process itself (Sanchez et al. 2020), e.g., different thermomechanical degradation history, batch-to-batch heterogeneity, or variations in filament quality.

Despite these differences, all batches exhibited stable load-bearing behavior under static loading, supporting the structural consistency, hence replicability, of the OS crutch across the tested distributed manufacturing contexts. This aspect was further investigated by comparing the mean curves for batch B01 with n=5 and n=10 specimens (B01b), where an additional five crutches were fabricated and assembled under identical conditions to the original set. Figure 9b compares the mean curves for B01 and B01b, showing similar trends in deformation behavior, load peak, and failure. Even if the standard deviation of B01b slightly increases along the mean curve, the overall similarity of the two curves confirms the repeatability of the replication process under the same material and fabrication conditions, for instance, when replicating different sets of crutches at different fabrication times but within the same distributed manufacturing context.

**Figure 9.**
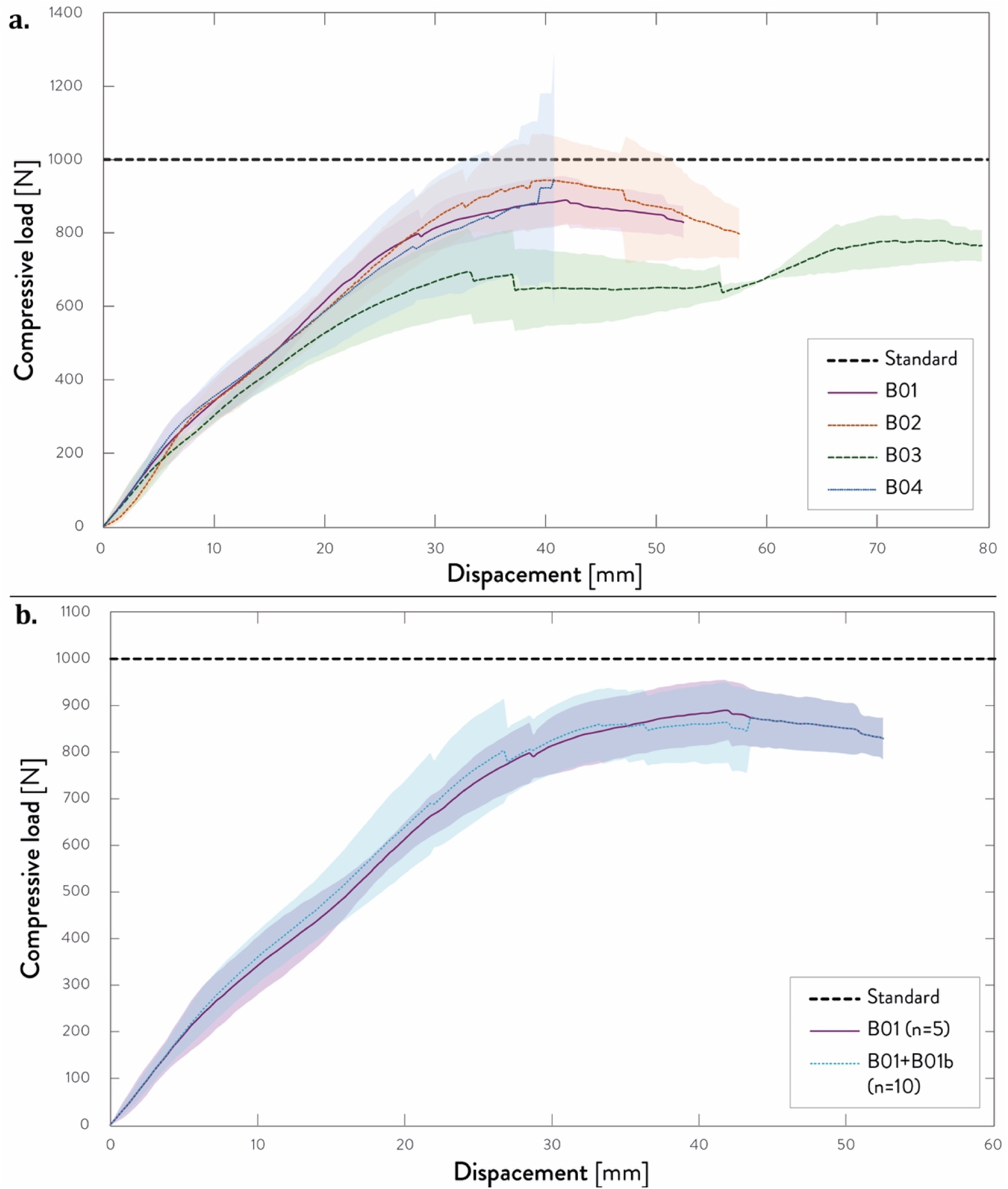
Mean compressive load versus displacement curves of: (a) the four batches of crutches; and (b) the first batch of n=5 and n=10 specimens (B01b). Shaded areas represent standard deviation from the mean curve, whereas the dotted lines represent the static load from standard ISO 11334-1:2007. The graphs compare the load-bearing capacity, deformation behavior, and variability across batches.

The static load threshold defined by the ISO standard, shown in Figure 11 and Figure S12 as dotted lines, indicates that only a subset of specimens exceeded the nominal static load of 1000 N, as confirmed by the mean maximum compressive loads of Table 3. Nevertheless, most crutches reached compressive loads at failure suitable for the bodyweight references (Colley et al. 2025), i.e., ∼900 N. These results support their intended purpose when used in pairs, as is common in most real-world applications of forearm crutches (Mottaghi et al. 2025). To this end, the safety factors for paired use (Table 3) represent how many multiples of 50% bodyweight each crutch would be expected to withstand before failure for representative male and female bodyweights (Colley et al. 2025), i.e., ∼1.86-2.22 and ∼2.23-2.66, respectively. Within the selected context, the design constraints set for an easily accessible, replicable, repairable, and adaptable assistive product result in a mechanical behavior that is consistent across the different simulated distributed manufacturing conditions.

**Table 3.** Mean experimental values and standard deviation from the mechanical static load tests, according to the different batches: maximum compressive loads, weights, and safety factors (single-use and use in pairs).

| Batch N. | Maximum compressive load (N) | Maximum weight (kg) | Safety factor (male, single-crutch) (-) | Safety factor (female, single-crutch) (-) | Safety factor (male, pairs) (-) | Safety factor (female, pairs) (-) |
| --- | --- | --- | --- | --- | --- | --- |
| B01 (n=5) | 893.4 ± 56.3 | 91.1 ± 5.7 | 1.05 ± 0.07 | 1.26 ± 0.08 | 2.11 ± 0.13 | 2.53 ± 0.16 |
| B01b (n=10) | 906.2 ± 80.4 | 92.8 ± 8.3 | 1.07 ± 0.10 | 1.29 ± 0.11 | 2.15 ± 0.19 | 2.57 ± 0.23 |
| B02 | 939.2 ± 99.8 | 95.7 ± 10.2 | 1.11 ± 0.12 | 1.33 ± 0.14 | 2.22 ± 0.24 | 2.66 ± 0.28 |
| B03 | 790.1 ± 32.0 | 80.5 ± 3.3 | 0.93 ± 0.04 | 1.12 ± 0.05 | 1.86 ± 0.08 | 2.23 ± 0.09 |
| B04 | 890.6 ± 175.0 | 90.8 ± 17.8 | 1.05 ± 0.21 | 1.26 ± 0.25 | 2.10 ± 0.41 | 2.52 ± 0.49 |

The failure mechanisms observed during the tests are shown in Figure 10, with additional insight into the specific failure points provided in Figure S12 (Supplementary Information, Section S3). In most cases, failure occurred in the wooden dowels, and it was mainly localized near the fastener regions connecting the handle to the upper dowels, where stress concentrations are expected due to load transfer at the interface between the parts. Some of these failures occurred without a complete or visible separation of the dowels, resulting in partial fractures or localized cracking. Only a limited number of crutches failed in the 3D-printed parts, i.e., 3 out of 25 samples, showing 3D-printing delamination parallel to the loading direction or cracking from the presence of interlayer voids. As visible from the tests, the failure shifts from custom 3D-printed components to standard parts that are accessible and easily available in low-resource contexts, prioritizing easily replaceable components in distributed assistive technology scenarios. On the one hand, this aspect simplifies the repair, replacement, adaptation, or reuse of the crutches with minimal tools and technical expertise (Antoniou et al. 2021), supporting circular design strategies (Kane et al. 2018; Romani et al. 2023) by replacing the shorter dowel. On the other hand, the wooden parts introduce additional variability in the mechanical behavior of the crutches, according to the intrinsic heterogeneity of the material that can be faced in distributed manufacturing contexts. In summary, the results confirm that the different replication settings keep functional equivalence across most of the tested batches, showing similar load behavior and failure modes despite differences in the maximum load peaks. It also confirms the viability of the design approach in considering replicability as one of the key aspects for distributed assistive technology.

**Figure 10.**
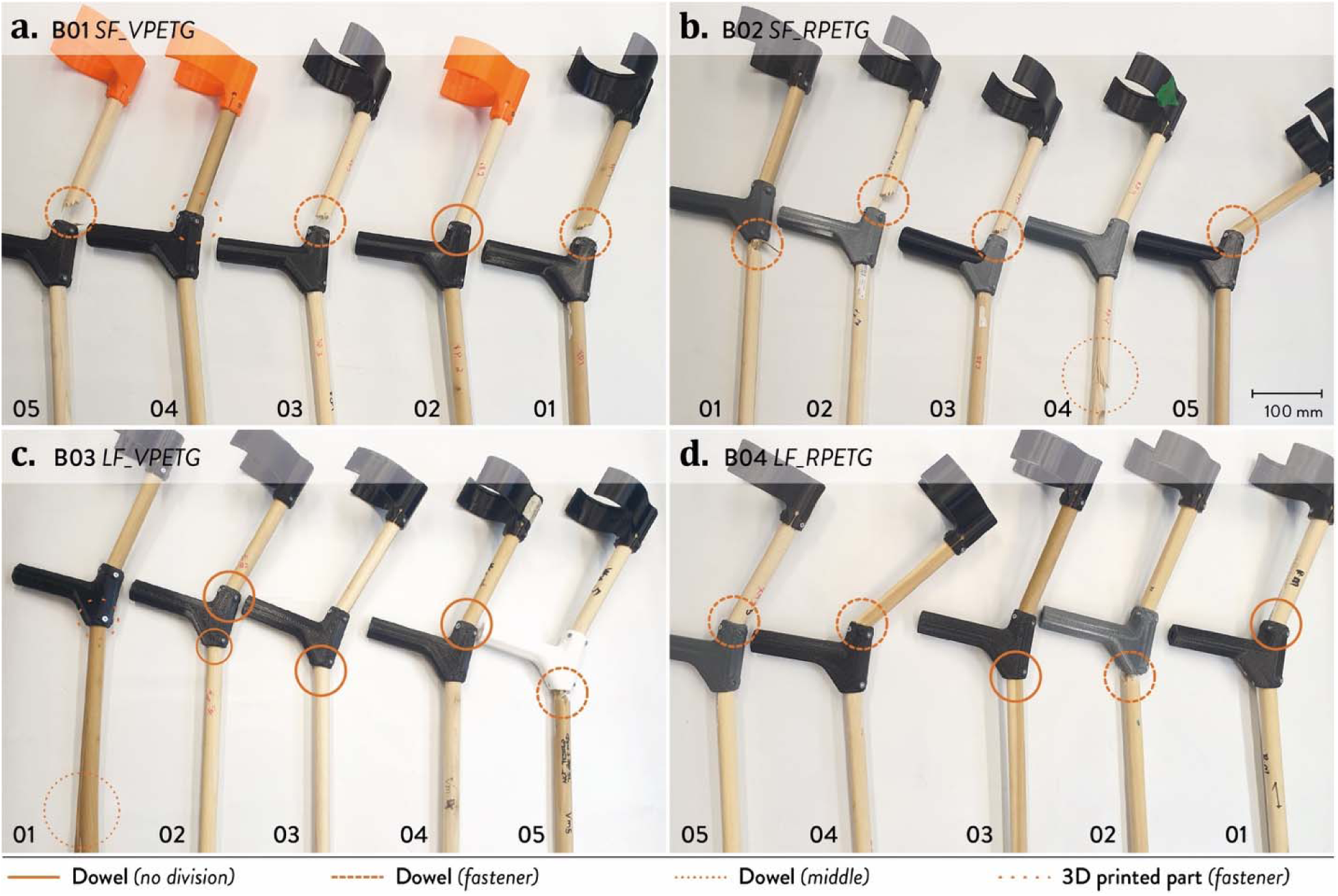
Failure points and mechanisms observed in the tested crutches according to the specific batch: (a) B01; (b) B02; (c) B03; and (d) B04. Each crutch is marked in its failure point with a line pattern according to the different failure mechanisms (continuous: dowel failure without division; dashed: dowel failure located at fasteners; dotted: dowel failure in a middle cross-section; loosely dotted: 3D printed part failure located at fasteners).

### Economic analysis

After evaluating the replicability of the crutches from a functional perspective, an economic analysis was conducted on the different batches to understand the variation in the affordability of the OS crutch introduced by the simulated distributed manufacturing context considered in the previous experimental steps. Cost calculations for the 3D-printed parts and assembled crutches for each batch are available in the Supplementary Information (Section S4) and on OSF (Romani et al. 2026). According to Table 4, the total cost of a single crutch ranged between 16.49 USD (B01) and 19.78 USD (B04), which means ∼33-40 USD for a set of two crutches. Despite the changes in the material feedstock origin and 3D printing scale, the cost difference is limited among the batches, i.e., within ∼3.30 USD per crutch, supporting a feasible replication of the same design without large cost changes.

**Table 4.** Costs of the different types of parts according to the different batches, showing the absolute and percentage costs for the 3D printed parts made of PETG, TPU, wooden dowels, and fasteners, together with the total costs for a single crutch and a set of two.

| Parts | B01 |  | B02 |  | B03 |  | B04 |  |
| --- | --- | --- | --- | --- | --- | --- | --- | --- |
|  | USD | % | USD | % | USD | % | USD | % |
| PETG parts | 5.48 | 33.2 | 7.42 | 40.2 | 6.84 | 38.3 | 8.77 | 44.3 |
| TPU parts | 4.11 | 24.9 | 4.11 | 22.3 | 4.11 | 23.0 | 4.11 | 20.8 |
| Dowels | 5.98 | 36.3 | 5.98 | 32.5 | 5.98 | 33.5 | 5.98 | 30.2 |
| Fasteners | 0.92 | 5.6 | 0.92 | 5.0 | 0.92 | 5.2 | 0.92 | 4.7 |
| **Total (x1 crutch)** | **16.49** | **//** | **18.44** | **//** | **17.85** | **//** | **19.78** | **//** |
| **Total (x2 crutches)** | **32.98** | **//** | **36.88** | **//** | **35.71** | **//** | **39.55** | **//** |

This fact is also confirmed by the percentage costs of the 3D printed components and the required off-the-shelf components. Across all batches, costs are mainly influenced by structural components, with PETG parts and wooden dowels contributing most to cost variation. PETG parts account for up to ∼44% of the costs, whereas dowels represent 30-36% of the total, depending on the batch. The increase in total cost across batches is mainly due to changes in the material feedstock rather than the scaling-up of fabrication. As visible in the detailed analysis in the Supplementary Information (Section S4), moving toward large-scale 3D printing of multiple components increases the energy costs due to higher power consumption of the system, e.g., for larger heating beds. This aspect, however, can be mitigated through alternative bed-adhesion strategies that do not require a heated bed.

These values are consistent with the design intention of using selected low-cost off-the-shelf components with custom, easily-replicable 3D-printable parts, enabling affordable replication of OS assistive products for the targeted use contexts with fewer constraints on the actual availability of off-the-shelf parts. Furthermore, the cost analysis shows that circular economy strategies to extend the product lifecycle are also feasible, as replacing, repairing, or adapting parts results in limited costs, e.g., ∼4-6 USD. Overall these results highlight that replicability-oriented design choices can ensure affordability of OS assistive products across different distributed manufacturing contexts without significant variations in costs.

### Replicability challenges in low-resource distributed manufacturing of OS products

Going beyond the specific case study, some key considerations can be made on replicability in OS products. First, it should not be treated as an aspect achieved only by formally meeting the requirements of OS licenses or to be evaluated at a later stage (Antoniou et al. 2021).

Replicability needs to be clearly considered from the early stages of OS product development, as part of the objectives and constraints considered in the design process. As suggested by Omer et al. (Omer et al. 2024), it requires a proactive approach toward Design for Distributed Manufacturing (DfDM), which considers the relevant variables affecting the replicability of the specific OS product, such as product function, target users, and the expected variability of local resources, components, fabrication settings, and builder’s expertise.

For the same reason, replicability issues in OSH development cannot be evaluated following a single, standardized protocol; instead, replicability assessment should be intentionally designed and included as a proper step of product development, ensuring coherence between the intended function of the product and its plausible local contexts. To this end, selecting appropriate evaluation methods to investigate replicability represents a key aspect, which should be guided by both the functional requirements of the product and its potential contexts of local replication, varying from case to case. In this work, replicability was explored by combining DfAM strategies, qualitative assessment of the fabrication and assembly quality, mechanical static load testing, and economic analysis. These aspects were selected to represent both the product application, i.e., mobility aid, and some relevant variables of the replication context, which means low-resource distributed manufacturing settings. Other relevant variables, such as user tests, were intentionally excluded at this stage, as they represent a following step in the evaluation, and are more focused on the product itself.

As shown by the comparison between the different batches, OSH must also be intentionally designed to tolerate some expected changes in the main variables related to its local fabrication. In other words, optimizing them for a single manufacturing setup should be avoided; instead, OSH products should intentionally accommodate the key variability from distributed manufacturing. For instance, this intention is made explicit in the failure mechanisms of the case study, where failures in the crutches are shifted from custom 3D printed parts to low-cost, widely available off-the-shelf components. This approach also simplifies repair, replacement, and adaptation for different users, supporting long-term adoption of this OSH product in low-resource distributed manufacturing settings, and can be seen as an example of an intentional design choice that supports the replicability of OS products, including assistive technology.

To sum up, this work shows how considering replicability as an early-stage design constraint, and keeping it during the product development process, helps include variability from local contexts into the design of OSH, or at least, recognize it as a factor influencing its real-world adoption.

### Limitations and future perspectives

While this work investigates replicability in OS products, especially assistive technology, from a practical perspective, it also shows several limitations and possible future directions. Focusing on the specific product, dynamic load tests are also recommended by the ISO standard to consider common actions performed with the crutches, e.g., walking or stair climbing. Although forearm crutches are used in pairs to effectively distribute the user’s weight across both supports, further testing and optimization are needed to assess performance under dynamic loading and improve results from static loading. Beyond specific functional aspects, replicability in this applicative context is also influenced by regulatory factors, which are still challenging for OS assistive products and differ across geographical contexts, e.g., different classifications and risk classes of medical devices.

From the replicability assessment, additional elements could also be explored to introduce other sources of variability, such as different types of dowels, self-made recycled filaments, different rigid materials, alternative options to replace TPU parts, second-hand parts, or different fastening options. Ongoing work is exploring different assembly solutions, including fastener-free connections and substituting flexible 3D-printed TPU with readily available or circular alternatives, e.g., components repurposed from discarded common products. Similarly, the study does not include geographic replication to better simulate a distributed manufacturing context. Future work should include geographical variables as a next step to deepen the evaluation of replicability and to consider them during the design process. As mentioned before, user tests and feedback were not included at this stage, although they provide key insights to improve the overall design. Future work will also include user tests (Wang et al. 2021), which could be conducted in specific distributed contexts or consider medium- and long-term use strategies, e.g., feedback on reuse, repair, and adaptation. Finally, even if representative, the study focuses on a single application context, and some aspects of replicability could have been omitted due to different requirements for specific OS assistive products.

## Conclusion

This work investigated replicability in OS assistive products in distributed manufacturing contexts through the design, fabrication, and experimental assessment of a two-piece OS forearm crutch. Within this study, replicability was included as a clear design objective and explored through controlled variables of plausible elements that change in distributed manufacturing contexts, such as material feedstock origins, 3D printing equipment, and fabrication strategies.

Focusing on the specific case study, the results show that the specific OS crutches can be fabricated using both small- and large-format FFF 3D printers, either with virgin or recycled PETG as rigid materials, ensuring a functional equivalence within the tested variations and broad accessibility to the product. Static load tests resulted in comparable load-bearing behavior across batches, e.g., from the mean average curves, reaching loads and safety factors suitable when the crutches are used in pairs, i.e., up to ∼940 N. Repeatability under identical conditions was further confirmed by the comparison between batches of n=5 and n=10 samples. The failure mechanisms were similar in most of the crutches and focused on the dowels, i.e., 22 out of 25 samples, shifting failure from custom 3D printed parts to low-cost and easily available components to facilitate product lifecycle extension in low-resource contexts, e.g., through maintenance, repair, and adaptation. The economic analysis further confirms limited cost variability across the batches, i.e., ∼16.50-20.00 USD each, with variations mostly due to the material feedstock, hence supporting the replication of the same design without significant cost variation. The OS crutches presented in this work are therefore a suitable option for people who need affordable, accessible mobility aid products in low-resource settings.

This work highlights the need to include replicability as a design-driven aspect from early-stage design and to consider it during the whole product development process. According to the results, OSH products, including assistive technology, should be intentionally designed to tolerate variability due to the local replication context, such as local resources, components, fabrication settings, and builder’s expertise, while ensuring comparable product functionality. To this end, replicability can be seen as a design constraint that influences the overall product development and guides real-adoption strategies, such as disassembly, repairability, repurpose, and adaptation for new users, supporting product lifecycle extension. Although the specific evaluation methods for its assessment may change according to the specific product and application context, this work demonstrates the relevance of designing specific approaches to assess replicability during the development of OS assistive products to support their real-world adoption in distributed manufacturing contexts.

## Data Availability

All data in the present work are available online at: https://doi.org/10.17605/OSF.IO/N8M5Y

## Acknowledgements

The authors would like to thank Morgan Woods for technical advice during the redesign of the testing rig and Anita So for the initial discussion about the redesign of the crutches.

## Additional information

Data are available in the Supporting Information and in the following OSF repository: https://osf.io/n8m5y/

## CRediT Author Statement

**Alessia Romani:** Conceptualization; Methodology; Software; Validation; Formal analysis; Investigation; Data curation; Writing – Original Draft; Writing – Review & Editing; Visualization; Supervision; Project Administration; **Rebecca Kaaya Nansubuga:** Formal analysis; Investigation; Data curation; Writing – original draft; Writing – Review & Editing; **Maryam Mottaghi:** Formal analysis; Investigation; Data curation; Writing – original draft; Writing – Review & Editing; **Danielle Munang:** Software; Formal analysis; Investigation; Data curation; Writing – Review & Editing; **Emily Bow Pearce:** Software; Investigation; Data curation; Writing – Review & Editing; **Pooja Viswanathan:** Validation; Writing – Review & Editing; Supervision; Funding Acquisition. **Thomas Jenkyn:** Validation; Writing – Review & Editing; Supervision; Funding Acquisition. **Tarek Loubani:** Validation; Writing – Review & Editing; Supervision; Funding Acquisition. **Jacob Reeves:** Conceptualization; Methodology; Software; Validation; Writing – Original draft; Writing – Review & Editing; Project administration; Funding Acquisition. **Joshua M. Pearce:** Conceptualization; Methodology; Validation; Resources; Writing – Original Draft; Writing – Review & Editing; Supervision; Project administration; Funding Acquisition.

