## Supplementary information for "Design for replicability in open-source distributed assistive technology for low-resource settings: a case study of two-piece 3D-printed forearm crutches"

This supplementary information document consists of three sub-sections. In detail:

- Sub-section S1 contains additional information about the assembly of the two-piece 3D-printed forearm crutch.
- Sub-section S2 contains further details about the design, manufacturing, and assembly of the open-source testing rig for static load testing.
- Sub-section S3 includes additional information about the results of the static load tests.
- Sub-section S4 presents further data about the economic analysis of the crutches.

### S1. Assembly of the crutches

This section provides the assembly instructions for the crutches. They are made of both custom and off-the-shelf parts. The former can be made with a desktop-size fused filament fabrication (FFF) 3D printer, whereas the latter can be easily purchased at hardware stores and cut to the desired length (for wooden dowels). As mentioned in the main text (Section "Design of the forearm crutch for low-resource distributed manufacturing contexts"), the assembly is made of three sub-assemblies: (i) cuff; (ii) handle; and (iii) crutch base. The components needed for the assembly are listed in Table S1. A detailed version of the bill of materials (BOM) is available on OSF, together with the 3D model of the assembly (Romani et al. 2026).

Table S1. BOM of the crutch: ID, part name, quantities, costs in USD, materials, and suppliers.

| Sub-assembly | Part | ID | Type | Q.ty | Cost per unit* | Material/Source |
|---|---|---|---|---|---|---|
| Cuff | Cuff body | C01 | Custom (FFF 3D printing) | 1 | 1.58-2.61 | Virgin or recycled PETG (ELEGOO US 2026; Spectrum Filaments 2022) |
| | Cuff grip | C02 | Custom (FFF 3D printing) | 1 | 1.38 | TPU 90 Shore A (PolyMaker 2026) |
| | Upper dowel | C03 | Off-the-shelf | 1 | 1.20 | Hardwood (Alexandria Moulding, The Home Depot) |
| Handle | Handle body | H01 | Custom (FFF 3D printing) | 1 | 3.04-4.74 | Virgin or recycled PETG (ELEGOO US 2026; Spectrum Filaments 2022) |
| | Handle grip | H02* | Custom (FFF 3D printing) | 1 | 1.47 | TPU 90 Shore A (PolyMaker 2026) |
| Base | Lower dowel | B01 | Off-the-shelf | 1 | 4.79 | Hardwood (Alexandria Moulding, The Home Depot) |
| | Washer | B02 | Custom (FFF 3D printing) | 1 | 0.06 | TPU 90 Shore A (PolyMaker 2026) |
| | Foot body | B03* | Custom (FFF 3D printing) | 1 | 0.50-0.84 | Virgin or recycled PETG (ELEGOO US 2026; Spectrum Filaments 2022) |
| | Living Joint | B04 | Custom (FFF 3D printing) | 1 | 0.60 | TPU 90 Shore A (PolyMaker 2026) |
| | Foot base | B05* | Custom (FFF 3D printing) | 1 | 0.35-0.57 | Virgin or recycled PETG (ELEGOO US 2026; Spectrum Filaments 2022) |
| | Foot base grip | B06 | Custom (FFF 3D printing) | 1 | 0.60 | TPU 90 Shore A (PolyMaker 2026) |
| Fastener | #6 x 5/8" flat head wood screws | F01 | Off-the-shelf | 13 | 0.07 | AISI 304 Stainless steel (DTGN, Amazon) |

**= Alternative 3D model for easy disassembly or fabrication available.*

The preliminary step involves fabricating the custom parts and purchasing the off-the-shelf components. The STL files for the 3D-printable parts are available in the OSF repository (Romani et al. 2026), along with the 3MF files containing the recommended 3D-printing settings. The parameters used for this work are shown in the main text (Section "Fabrication of the 3D printed components", Table 2). The parts are suitable for desktop-size FFF 3D printers. Off-the-shelf components consist of cylindrical hardwood dowels with 22.3 mm (7/8 inch) diameter and #6 x 5/8 inch flat-head wood screws (Table S1). The formers need to be cut to the desired lengths to obtain the upper and lower dowels (C03 and B01), which are 210 and 800 mm in this specific work, respectively. Their length can be customized according to the specific user's proportions by cutting shorter or longer dowels after checking the most appropriate measures. In general, the cuff should be 25-50 mm below the bend of the elbow, and the overall height of the crutch should allow the handle to be grasped at the wrist crease when the arm is extended (Bauer et al. 1991).

After fabricating and purchasing the required parts, the crutch can be assembled according to the following instructions. Some steps require additional tools, such as a permanent marker, medium-grit sandpaper, and an electric drill compatible with Phillips or square driver bits, depending on the selected screw head type. Appropriate personal protective equipment (PPE) must be worn during the assembly procedure, such as protective gloves and safety glasses. A protective mask should be used when sanding wooden dowels, and all drilling operations should be performed on parts properly fixed to working surfaces, e.g., using clamps.

- **Surface preparation of the hardwood dowels:** Before assembling the main parts, each end of the cut dowels (C03 and B01) must be sanded to remove sharp edges, splinters, or surface irregularities that could make the insertion into the 3D-printed components difficult or cause damage during handling. These portions of the dowels should be ideally smooth and slightly chamfered to facilitate insertion. In addition, the entire length of the dowels must be visually inspected for any surface irregularities that could harm the user, and such irregularities must be removed with medium-grit sandpaper. After sanding, any residual dust should be removed before continuing with the assembly.
- **Assembly of the forearm cuff:** The flexible cuff flat grip (C02) is fixed on the main body of the cuff (C01). After bending the flexible part, the embossed insert on the upper surface is aligned with the dedicated slot of the rigid cuff, then pushed into the slot (Figure S1a) and slid along one side (Figure S1b). The remaining part of the insert is finally pushed into the rigid slot by applying slight pressure on the flexible material (Figure S1c) and sliding it toward the end of the slot (Figure S1d). This action allows for a solid yet reversible assembly of the two components without the need for glue, as the flat grip naturally tends to unbend and lock into the rigid slot of the cuff. Once the cuff is ready, the upper dowel (C03) is inserted vertically into the cuff opening until it reaches the end of the cavity (Figure S1e). Full insertion is required to guarantee correct load transfer between the two components.

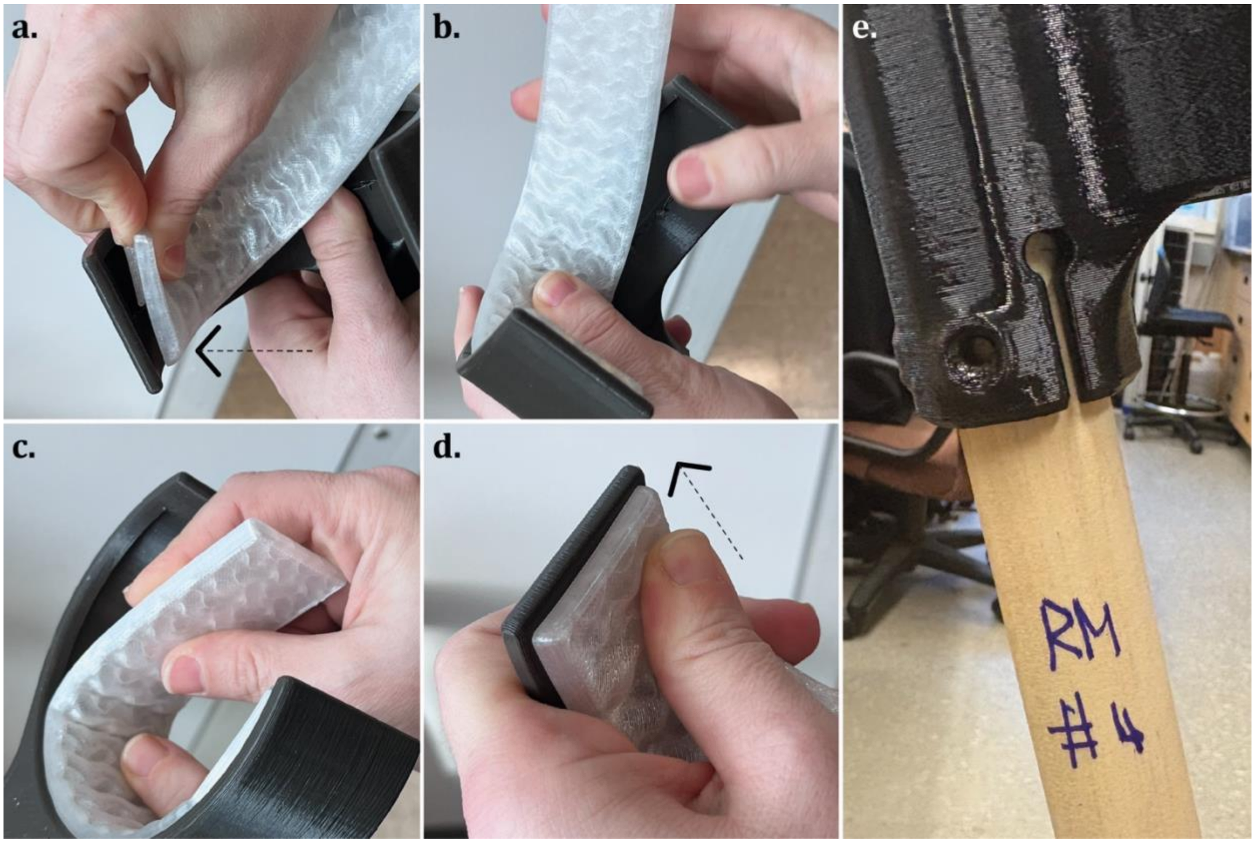


Figure S1. Assembly of the cuff grip and body: (a) starting insertion of the grip insert into the cuff slot; (b) insertion of the remaining portion of the grip; (c) sliding and pressure gesture to place the whole part; and (d) locking gesture to ensure proper assembly of the components. The upper dowel (e) can then be assembled into the cavity of the cuff body.

- **Assembly of the handle:** The flexible handle grip (H02) is fixed to the main body of the handle (H01) by sliding it along the rigid part until it fully reaches the end (Figure S2a). The free end of the upper dowel (C02) is inserted vertically into the four-screw cavity of the handle, ensuring its full insertion (Figure S2b).

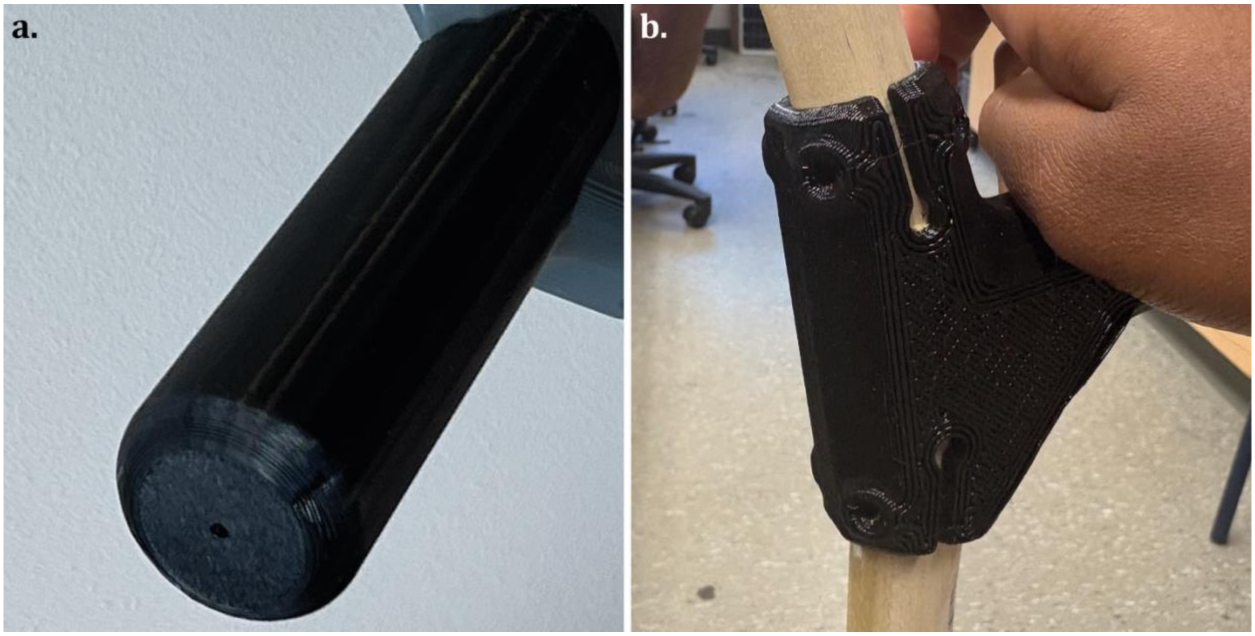


Figure S2. Assembly of the handle: (a) insertion of the handle grip on the handle body; and (b) insertion of the upper and lower dowels into the cavities of the handle.

- **Assembly of the foot:** The living joint (B04) is connected to the dedicated slot of the foot base (B05) by squeezing its lower insert and pushing it into the dedicated slot of the rigid component (Figure S3a). The upper insert of the living joint is then inserted into the lower slot of the foot body (B03), following the same procedure used to connect B05 and B04 (Figure S3b). The foot base grip (B06) is finally inserted into the lower slot of the foot base, using the same sliding and locking mechanism applied to the previous flexible parts of the foot (Figure S3c). Similar to the cuff, these steps ensure a solid yet reversible assembly of the parts without glue or additional fasteners, relying on the mechanical interlocking between the rigid and flexible parts (Figure S3d).

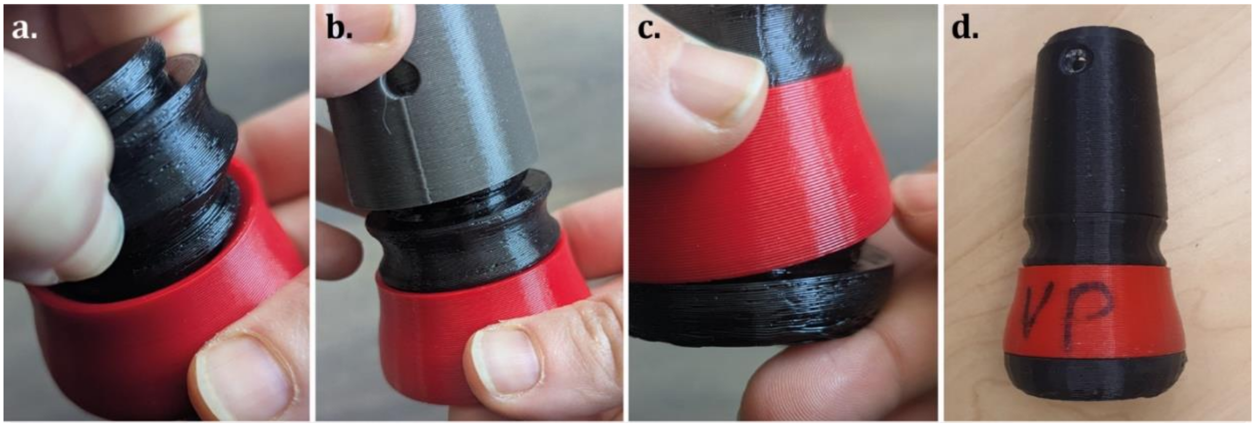


Figure S3. Assembly of the foot: (a) insertion of the living joint into the foot base and (b) into the foot body; (c) insertion of the foot grip into the foot base; and (d) assembled foot.

- **Assembly of the base:** The foot assembly, prepared in the previous step, is fixed to the opposite end of the lower dowel (B01) together with a flexible washer (B02) to ensure good fixation and load distribution (Figure S4a). In both cases, the lower dowel must be pushed in until it is fully inserted into the two 3D-printed cavities (Figure S4b).

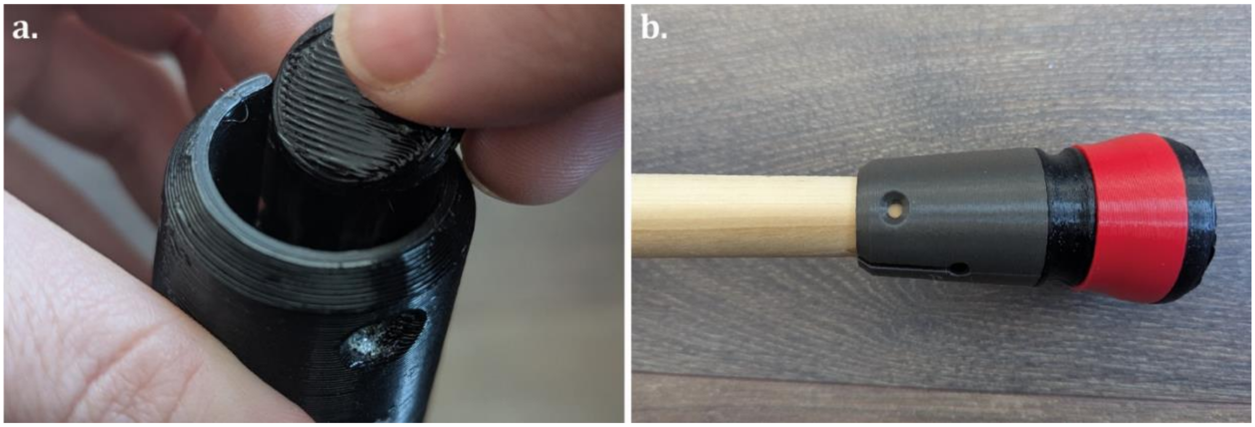


Figure S4. Assembly of the base: (a) insertion of the washer into the foot assembly; and (b) insertion of the lower dowel into the cavity of the foot body

- **Fit verification and component alignment:** Before drilling and fastening the components, the correct insertion and depth of the wooden dowels (C03 and B01) must be visually and manually checked to avoid structural problems and premature failure during use. The alignment of the cuff and handle must also be checked at this stage, ensuring linear alignment, avoiding angular offsets or misalignments. They can be checked by laying the assembly on a flat surface, such as a table or the floor, using the flat sides of the cuff (C01) and handle (H01) as references (Figure S5). Minor adjustments can be made by rotating the components and rechecking the alignment with the same approach. If needed, additional washers can be added to the 3D-printed cavities of the cuff and handle to further improve overall assembly stability.

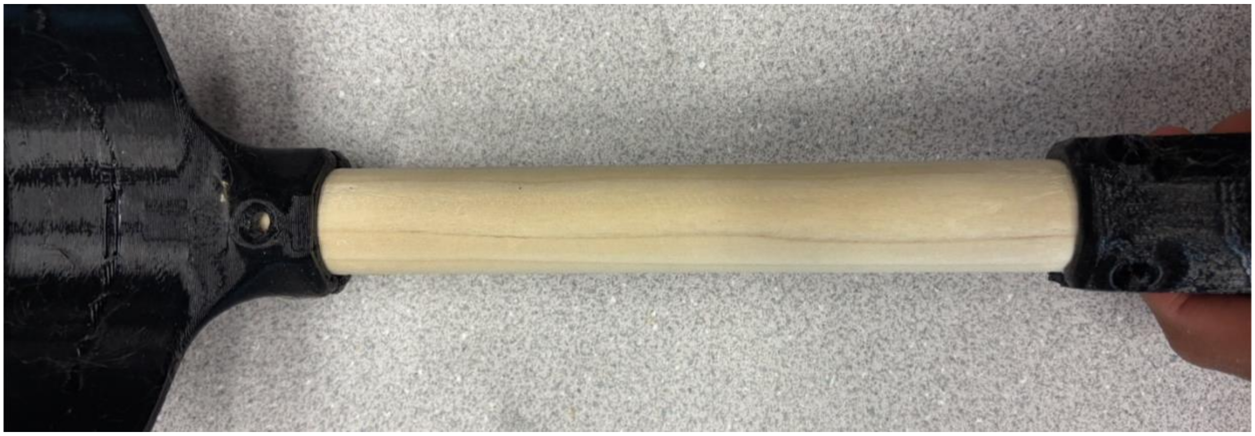

Figure S5. Checking the alignment of the 3D printed components with a reference flat surface.

- **Pilot hole drilling and screw placement:** Once the alignment is correct, pilot holes are drilled into the wooden dowels. The 3D-printed parts can be used as a positioning guide for drilling the holes, paying attention to avoid damage. As an alternative, they can be used to mark the hole positions with a permanent marker, removed for the drilling step, reassembled, and realigned before fastening. In both cases, an appropriate drill bit is recommended, e.g., 3.5 mm in this configuration, as well as clamping the components for precise results. Drilling must be performed carefully to avoid over-drilling or damaging components, as well as getting shorter or longer holes that can compromise the assembly or the structural integrity of the crutch during use. After drilling the holes, #6 x 5/8 inch flat-head wood screws (F01) with either square or Phillip head types are inserted (Figure S6a) and gradually tightened with an electric drill or a screwdriver, until securely fastened and avoiding over-tightening (Figures S6b and c). Wooden screws following imperial standards can be replaced with metric alternatives, in this case, M4 x 16 mm countersunk wood screws. This procedure is repeated until all 3D-printed components are properly secured to the dowels. The assembled crutch is shown in Figure S6d, and the second one can be assembled using the same procedure.

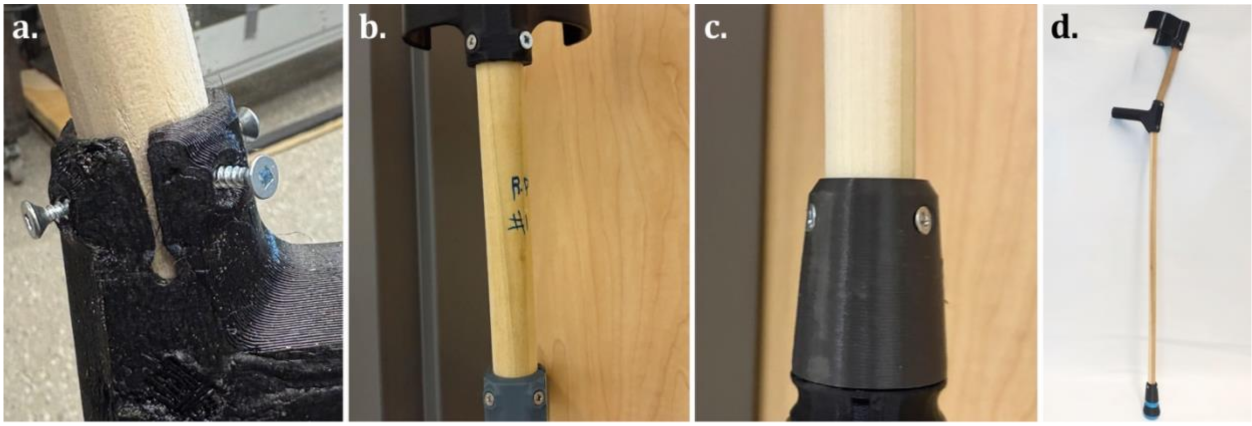


Figure S6. Fastening of the parts: (a) tightening of the wood screws into the pilot holes; (b and c) final tightening of the 3D printed parts; and (d) final assembled crutch.

All fasteners should be periodically checked to ensure that they are not loose and, if needed, retightened before use. In case of breaks or wear, the parts can be easily disassembled to allow for replacements, repair, or repurposing, e.g., adapting the length of the dowels to new users or changing any damaged component. The fasteners can be removed by using an electric drill or a manual screwdriver. The flexible 3D-printed components can be removed by squeezing and pressing them during removal, as during assembly.

Alternative 3D models of the rigid components of the foot are available on OSF (Romani et al. 2026) to simplify disassembly with a cylindrical stick or tool, such as a screwdriver or pen, or to facilitate the fabrication with TPU.

Figure S7 shows the removal of the flexible components of the foot from the alternative versions of the rigid parts, using a common screwdriver to simplify disassembly and avoid manual squeezing, hence reducing stress on hands. The lateral hole on the modified rigid parts allows the user to insert a screwdriver and use it as a lever to remove the flexible parts (Figure S7a). This approach can be used for all connections within the foot, i.e., between the foot base grip (B06) and the foot base (B05, Figure S7a), between the living joint (B04) and the foot base (Figure S7b), or between the living joint and the foot body (B03, Figure S7c).

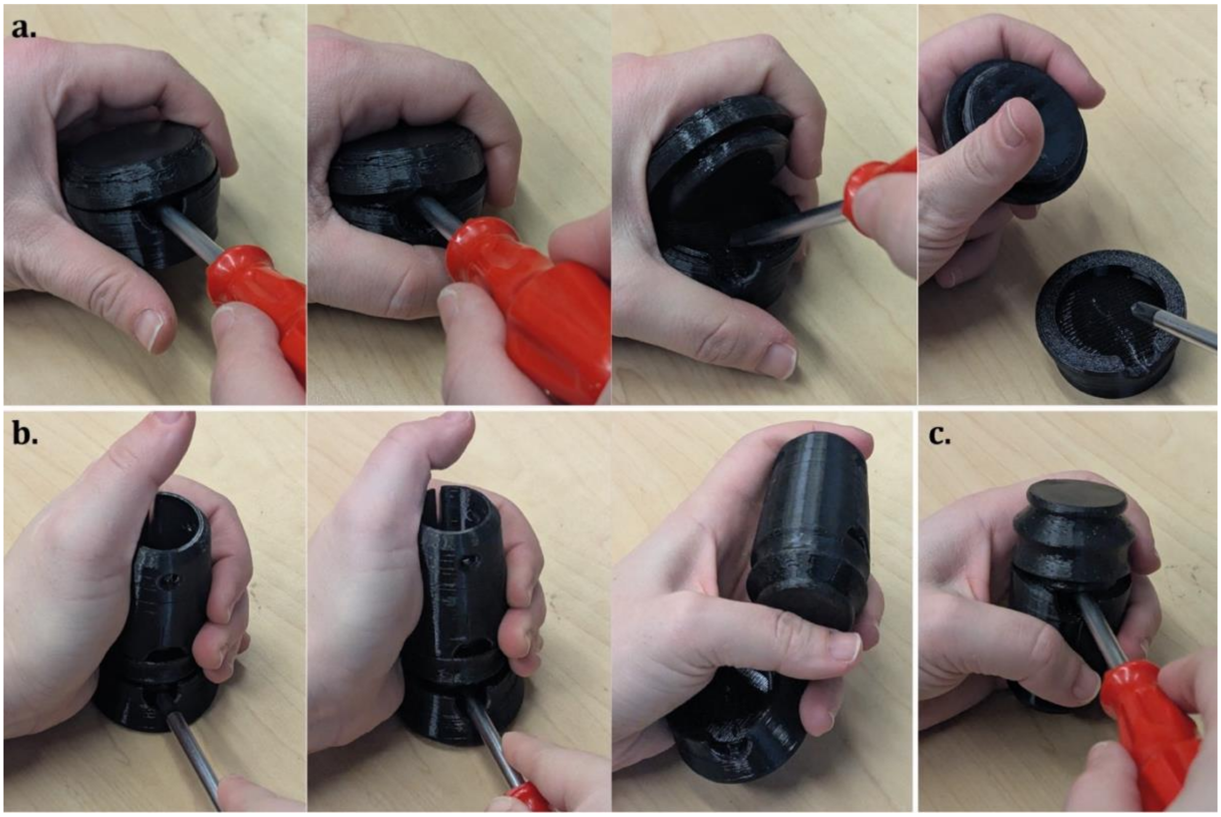


Figure S7. Disassembly of the alternative foot components: (a) removal of the foot base and (b, c) of the living joint with a screwdriver.

A flat version of the handle grip (H02, Figure S8) is available as an alternative part for easy fabrication and customization of the grip. After 3D printing, the grip can be assembled by bending its main body and connecting the lateral flange to the corresponding holes before inserting it on the handle (H01, Figure S8a). The two frontal pins can be inserted into the engraved hole of the handle and then secured through the missing flanges (Figure S8b) to block the grip before use (Figure S8c).

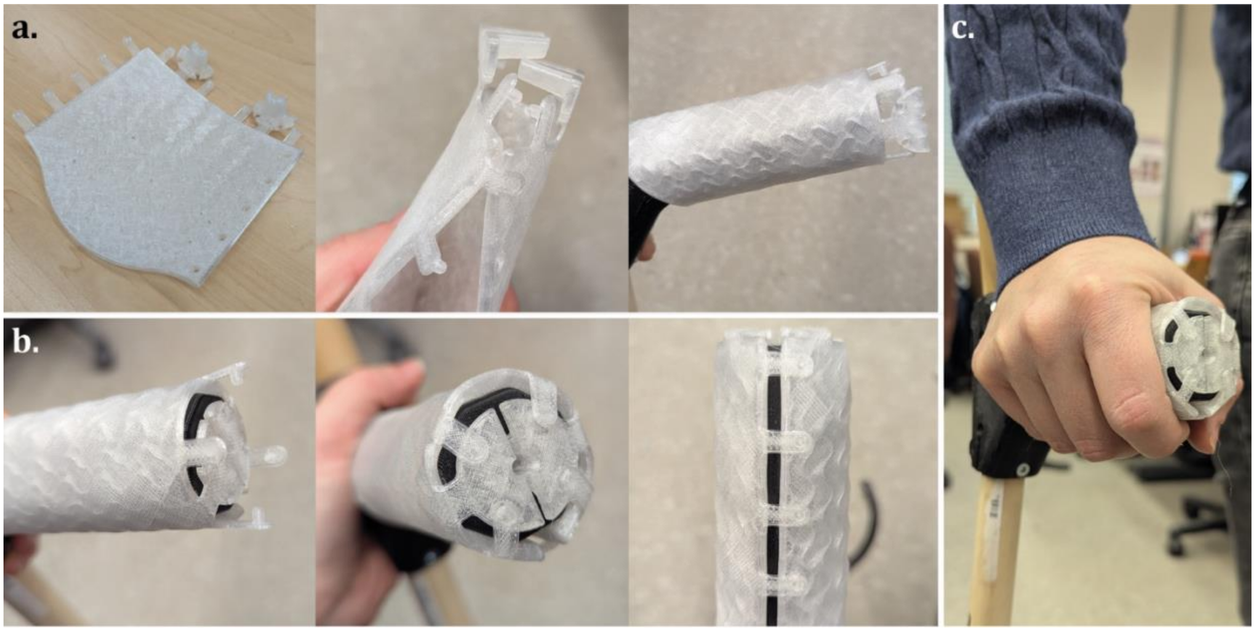


Figure S8. Assembly of the alternative flat handle grip: (a) removal of the foot base and (b, c) of the living joint with a screwdriver.

### S2. Design, manufacturing, and assembly of the testing rig

The crutch testing rig was made from custom 3D-printed and off-the-shelf components, modified where necessary to suit the purpose. The current version is a modification of the original design used by Mottaghi et al (Mottaghi et al. 2025), previously shared on OSF (Pearce et al. 2024), and is designed to fit a forearm crutch for static load testing according to the ISO 11334-1:2007 standard (ISO 2022). All design files (STEP files) are available on the OSF repository of this project (Romani et al. 2026).

Figure S9 shows the assembly of the testing rig during the test (on the left) and an exploded view of its main components (on the right). The BOM is visible in Table S2.

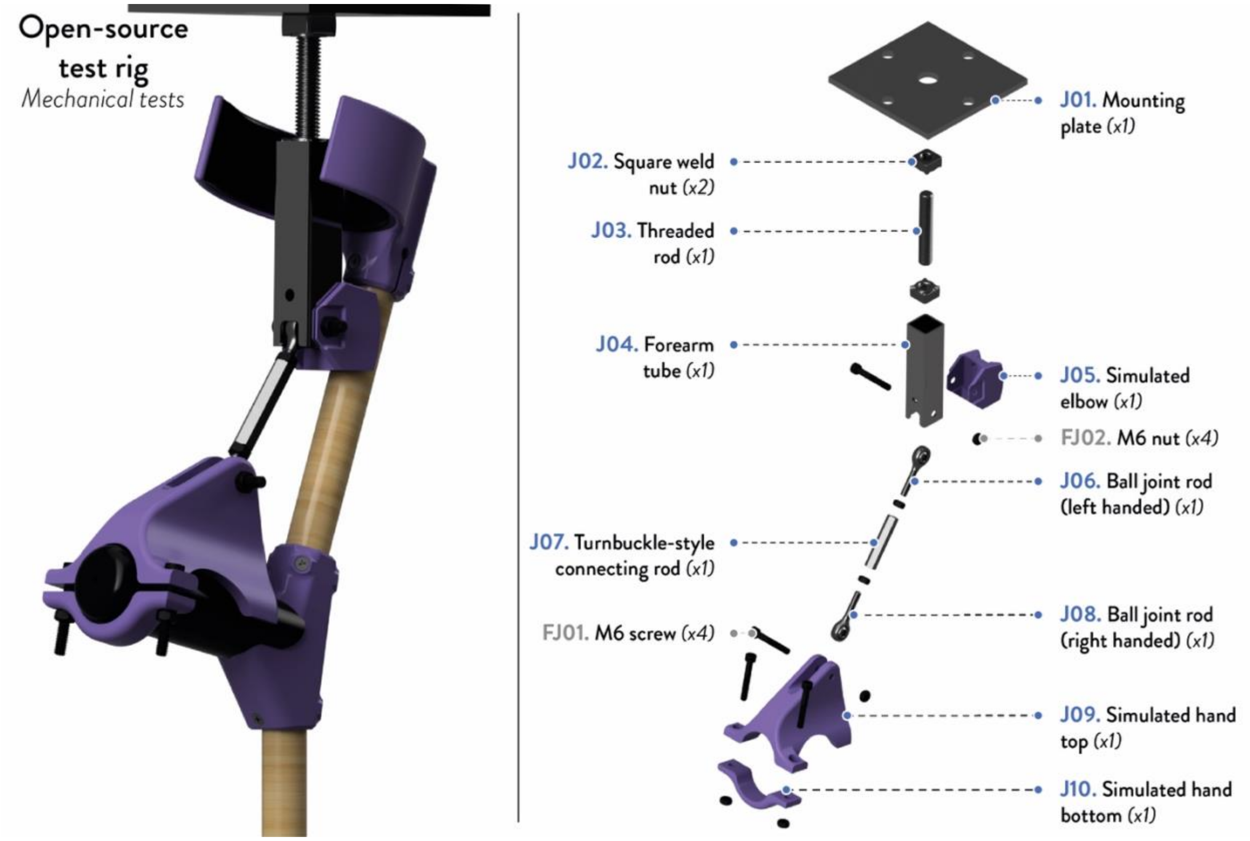


Figure S9. Preview of the assembled testing rig during use (on the left) and exploded view of the assembly, showing the position and quantities of the components (on the right).

Table S2. BOM of the testing rig, showing the ID, part name, quantities, costs in USD, materials, and suppliers.

| Part | ID | Type | Q.ty | Cost per unit* | Material | Source |
|---|---|---|---|---|---|---|
| Mounting plate | J01 | Off-the-shelf (drilling) | 1 | 23.08 | Mild steel | (Metal Supermarkets 2024) |
| 3/8 inch - 16 Threaded Square Weld Nut, 0.062 inch | J02 | Off-the-shelf (welding) | 2 | 0.52 | Steel | (McMaster-Carr 2026) |
| 3/8 inch - 16 Threaded Rod, 3 inch long | J03 | Off-the-shelf | 1 | 1.57 | Steel | (McMaster-Carr 2026) |
| Forearm tube | J04 | Off-the-shelf (drilling, cutting) | 1 | 2.92 | Mild steel | (Metal Supermarkets 2024) |
| Simulated elbow | J05 | Custom (FFF 3D printing) | 1 | 0.44 | PETG | (ELEGOO US 2026) |
| 1/4 inch - 28 Threaded Ball joint rod end with nut (left handed) | J06 | Off-the-shelf | 1 | 5.28 | Zinc-Plated Carbon steel | (McMaster-Carr 2026) |
| 1/4 inch - 28 Internal Turnbuckle-style connecting rod, 2 inch long | J07 | Off-the-shelf | 1 | 12.49 | Carbon steel | (McMaster-Carr 2026) |
| 1/4 inch - 28 Threaded Ball joint rod end with nut (right handed) | J08 | Off-the-shelf | 1 | 5.28 | Zinc-Plated Carbon steel | (McMaster-Carr 2026) |
| Simulated hand (top) | J09 | Custom (FFF 3D printing) | 1 | 2.51 | PETG | (ELEGOO US 2026) |
| Simulated hand (bottom) | J10 | Custom (FFF 3D printing) | 1 | 0.32 | PETG | (ELEGOO US 2026) |
| M6 x1 mm Stainless steel socket head screw, 40 mm long | FJ01 | Off-the-shelf | 4 | 0.25 | 18-8 Stainless steel | (McMaster-Carr 2026) |
| M6 x 1 mm zinc-plated steel hex nut | FJ02 | Off-the-shelf | 4 | 0.02 | Zinc-Plated steel | (McMaster-Carr 2026) |

**= Machining excluded (drilling, cutting, welding).*

#### *S2.1 3D printed components*

The 3D-printed components were made with Elegoo Rapid PETG filament (Shenzhen, China) (ELEGOO US 2026) on a Prusa MK3S (Prusa Research, Prague, Czech Republic), a desktop open-source RepRap printer, using a 0.8 mm nozzle. The gcode files were made with the open-source slicing software PrusaSlicer (Prusa3D by Josef Prusa 2026).

A gyroid infill pattern with 99% infill density, four wall perimeters, and a layer height of 0.4 mm were selected as the main parameters. The bed and nozzle 3D printing temperatures were set to

85 °C and 230 °C to ensure good bed adhesion. Support structures were used where necessary. The STL, gcode, and 3mf files of the 3D-printed components are available in the OSF repository (Romani et al. 2026).

#### *S2.2 Off-the-shelf components*

The off-the-shelf hardware components used to build the open-source crutch testing rig were primarily made of steel (BOM, Table S2). Four holes with a 19.05 mm (0.75 inch) diameter, radially located 127 mm (5 inches) apart, were drilled through the top mounting plate to allow attachment to a vertical hydraulic machine that provided the compressive force to test the crutches. The holes and dimensions of the mounting plate can be adjusted according to the dimensions of the specific plate on the hydraulic press used for the tests. Similarly, the forearm tube was drilled to allow attachment to the simulated elbow. Square weld nuts were welded to one end of the forearm tube and to the top mounting plate, allowing the pieces to be connected with a threaded rod. The ball joint rod ends were purposely selected with a swivel range that exceeds the 15-degree minimum requirement by the ISO 11334-1:2007 standard for the forearm to rotate in all directions (ISO 2022).

In the initial design (Mottaghi et al. 2025), the vertical compressive force was transferred to the crutches via a simulated forearm consisting of a forearm tube welded to two weld nuts at either end. This simulated forearm was replaced with a turnbuckle-style connecting rod to improve the replicability of the rig design and enable customization of the distance between the simulated elbow and hand, e.g., with different sizes and types of tested crutches.

#### *S2.3 Building and use instructions*

With the testing rig components prepared according to the required specifications, its building can be better achieved using additional tools such as an adjustable wrench or spanner and corresponding Allen keys fitting the screws. The testing rig can then be assembled using the following instructions.

- **Building the testing rig:** The mounting plate (J01) is connected to the forearm tube (J04) via a threaded rod (J03) threaded into square weld nuts (J02) welded to the mounting plate and to the forearm tube. The forearm tube is then mounted onto the simulated elbow (J05) such that the holes align. Both of these are then connected to a ball joint rod end (J06) and held together using an M6 screw (FJ01) and its corresponding nut (FJ02), such that this simulates the human elbow joint. The ball joint rod end (J06) is fitted to a turnbuckle-style connecting rod (J07) whose opposite end is attached to another ball joint end rod (J08). The ball joint of J08 is connected to the simulated hand top using an M6 screw and nut, simulating the human wrist joint. The simulated hand top (J09) is then connected to the simulated hand bottom (J10) using an M6 screw and nut, whose tightness can be adjusted to simulate the human hand gripping the crutch handle. The complete build should correspond to Figure S9.
- **Mounting the testing rig onto the hydraulic press:** The fully assembled testing rig is connected to a vertical hydraulic press, through which the compressive load is supplied via the holes in the mounting plate. The mounting plate is fixed in place using two 19.05 mm (0.75 inch) diameter hex screws (McMaster-Carr 2026). Please note that the two hex screws used to mount the rig to the hydraulic press are not included in the BOM (Table S2), as their specific model and dimensions vary depending on the press used for the

tests. For this reason, the holes of the mounting plate can be drilled according to the specifics of the testing press, and the suitable matching screws can vary accordingly.

- **Using the testing rig:** A fully assembled crutch is fixed to the testing rig such that the simulated elbow rests against the upper dowel (C03) and the simulated hand top and bottom grip the handle grip (H02), as shown in Figure S10a. The foot of the crutch is placed such that it is vertically below the loading direction (Figure S10b) to ensure that the load point mimics the force exerted on the crutch by the user during actual use. A laser, pointed temporarily vertically from the forearm tube, can be used to locate the appropriate foot position on the ground. After fixing the crutch, the position of the hydraulic press can be slightly adjusted to block the position of the crutch, paying attention to avoid applying any loads before the test (Figure S10c). To switch from one crutch to another during testing, loosen one of the M6 screws holding the simulated hand top and bottom together.

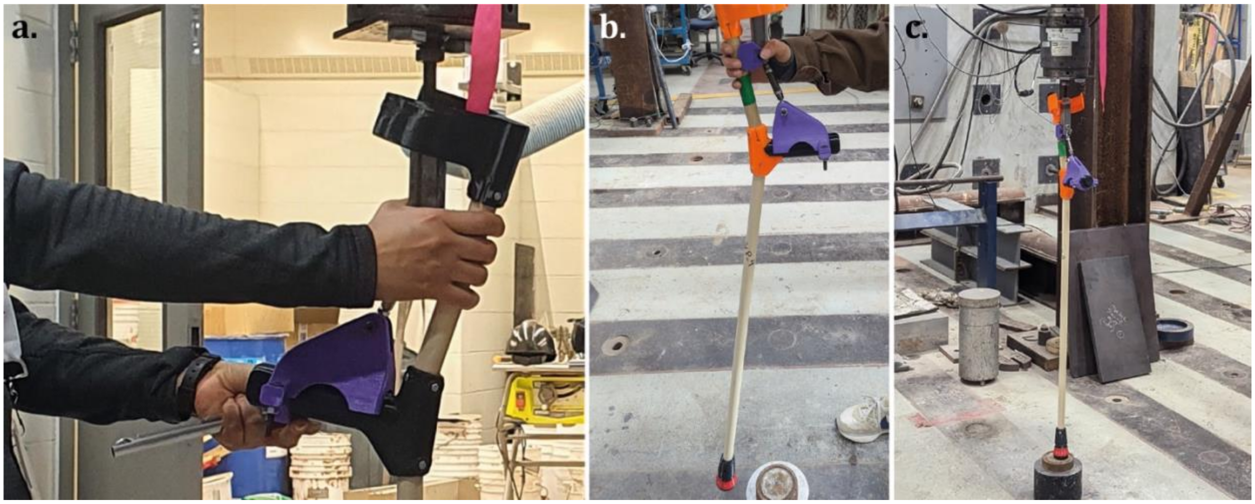


Figure S10. Use of the testing rig: (a) positioning of the simulated elbow against the upper dowel and the simulated hand top and bottom on the handle grip; (b) positioning of the crutch foot on the ground; and (c) well-positioned crutch ready for testing.

Depending on the height of the crutches being tested or the lowest height adjustment limit of the hydraulic press, solid blocks can be added on the ground before positioning the crutch for testing, or the testing rig itself can be lengthened. This can be achieved by adjusting the height of the threaded rod or the ball joint rod ends within the connecting tube until they reach their maximum length. A connecting rod that is left-hand threaded on one end and right-hand threaded on the other end, with corresponding left and right-hand threaded ball joint rod ends, was used to allow easier adjustment.

#### *S2.4 Costs of the testing rig*

The spreadsheet with the cost calculation is available on OSF. The analysis methodology is explained in the main text (Section "Economic analysis"). Table S3 shows the cost analysis of the 3D-printed parts, whereas Table S4 includes the total cost of the testing rig. For the sake of this work, only material costs were considered for off-the-shelf components requiring further machining, e.g., drilling, welding, cutting, assuming the internal availability of these fabrication methods.

Table S3. Nominal weight and 3D printing times of the rig parts made with virgin PETG.

| **Part** | **ID** | **Q.ty** | **Weight (g)** | **Time (min)** | **Costs** | |
|---|---|---|---|---|---|---|
| | | | | | **Elements** | **Per unit** |
| Simulated elbow | J05 | 1 | 32 | 56 | Material | 0.43 |
| | | | | | Energy | 0.01 |
| | | | | | Total | 0.44 |
| Simulated hand (top) | J09 | 1 | 181 | 290 | Material | 2.48 |
| | | | | | Energy | 0.03 |
| | | | | | Total | 2.51 |
| Simulated hand (bottom) | J10 | 1 | 23 | 42 | Material | 0.32 |
| | | | | | Energy | >0.01 |
| | | | | | Total | 0.32 |
| Total | | | 236 | 388 | // | 3.27 |

Table S4. BOM of the testing rig, showing the ID, part name, quantities, costs in USD, materials, and suppliers.

| **Part** | **ID** | **Type** | **Q.ty** | **Cost per unit*** | **Total cost*** |
|---|---|---|---|---|---|
| Mounting plate | J01 | Off-the-shelf | 1 | 23.08 | 23.08 |
| 3/8 inch - 16 Threaded Square Weld Nut, 0.062 inch | J02 | Off-the-shelf | 2 | 0.52 | 1.03 |
| 3/8 inch - 16 Threaded Rod, 3 inch long | J03 | Off-the-shelf | 1 | 1.57 | 1.57 |
| Forearm tube | J04 | Off-the-shelf | 1 | 2.92 | 2.92 |
| Simulated elbow | J05 | Custom | 1 | 0.44 | 0.44 |
| 1/4 inch - 28 Threaded Ball joint rod end with nut (left handed) | J06 | Off-the-shelf | 1 | 5.28 | 5.28 |
| 1/4 inch - 28 Internal Turnbuckle-style connecting rod, 2 inch long | J07 | Off-the-shelf | 1 | 12.49 | 12.49 |
| 1/4 inch - 28 Threaded Ball joint rod end with nut (right handed) | J08 | Off-the-shelf | 1 | 5.28 | 5.28 |
| Simulated hand (top) | J09 | Custom | 1 | 2.51 | 2.51 |
| Simulated hand (bottom) | J10 | Custom | 1 | 0.32 | 0.32 |
| M6 x1 mm Stainless steel socket head screw, 40 mm long | FJ01 | Off-the-shelf | 4 | 0.25 | 1.01 |
| M6 x 1 mm zinc-plated steel hex nut | FJ02 | Off-the-shelf | 4 | 0.02 | 0.09 |
| Total | | | | | 56.04 |

**= Machining excluded (drilling, cutting, welding).*

### S3. Additional information about the static load tests

The curves from the static load tests are shown in Figure S11, grouped by batch (B01, B02, B03, and B04). They were used to calculate the average curves presented in the main text (Section "Mechanical static load tests") and correspond to the visualization of data after displacement normalization and interpolation. For B01 (Figure S11a), the curves were divided according to the first or second batch of tested crutches (B01b, n=10), represented by the continuous and dotted curves, respectively. The datasheet with the full dataset and analysis of the static load tests is available on OSF (Romani et al. 2026).

Figure S12 shows the observed failure points for each crutch, also divided by batch and including the additional pictures of batch B01b (Figure S12a, Samples 6-10).

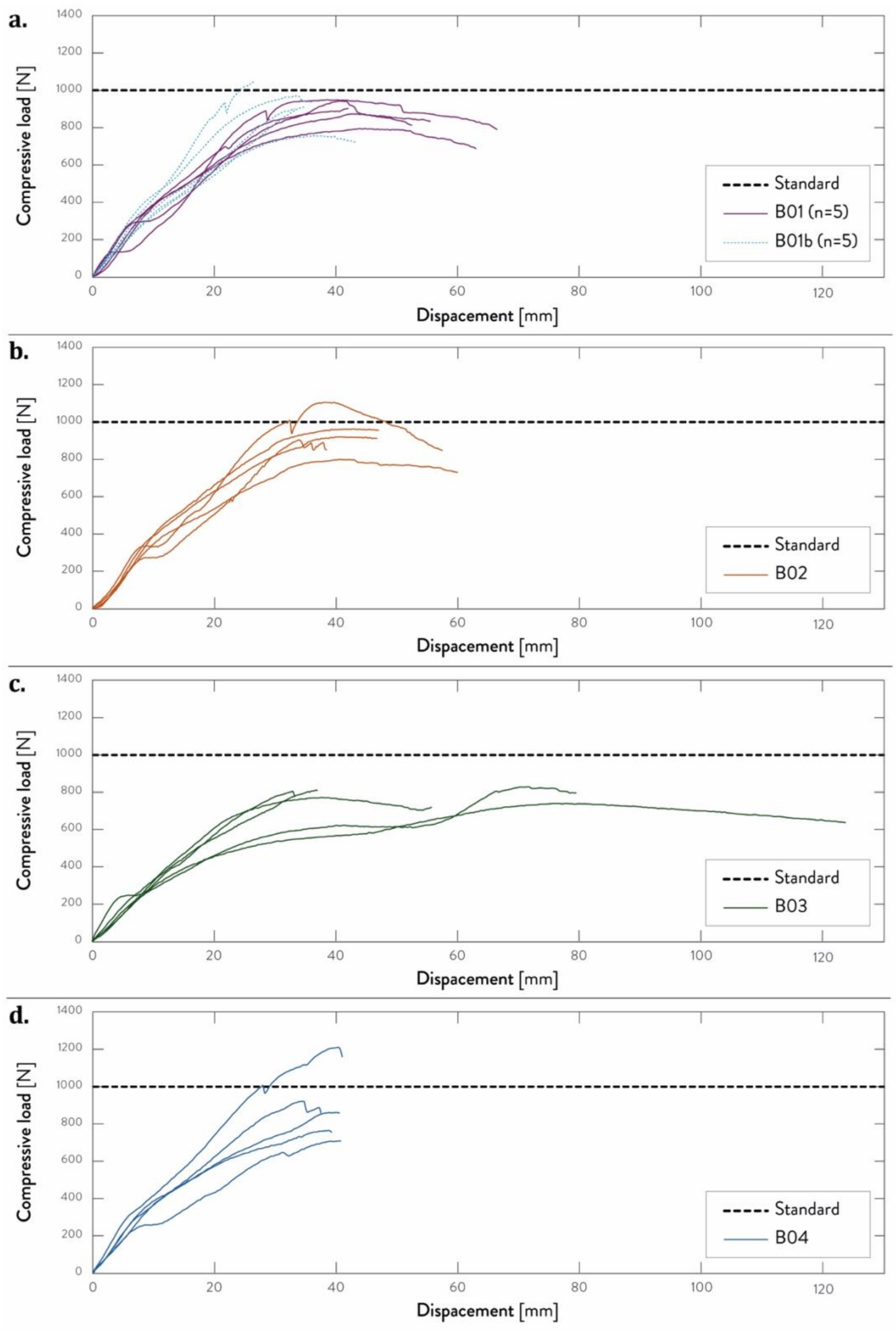


Figure S11. Single curves (compressive load versus displacement) of (a) B01 and B01b; (b) B02; (c) B03; and (d) B04. The dotted line represents the static load from standard ISO 11334-1:2007.

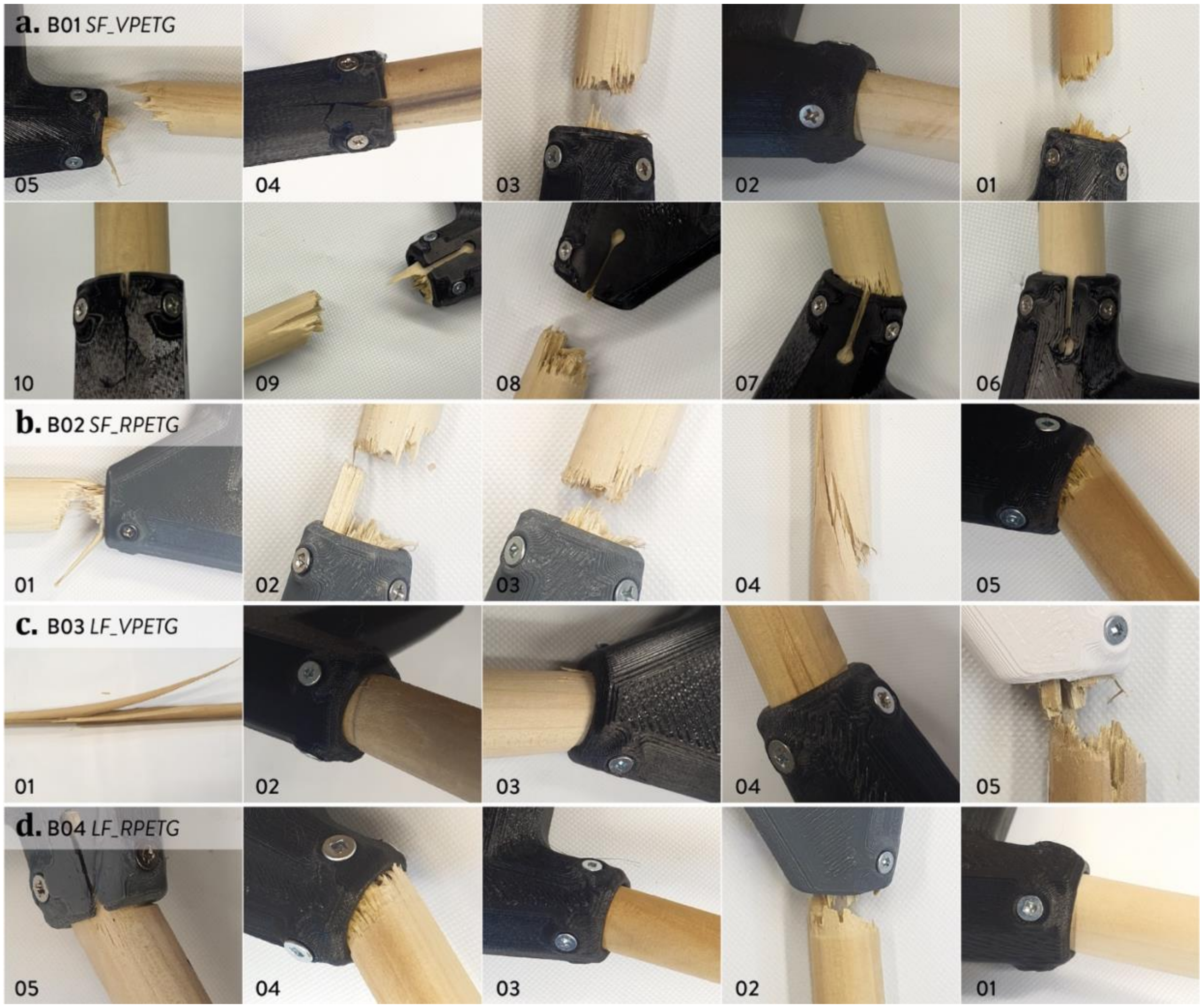


Figure S12. Insights of the failure points observed in the tested crutches, divided according to the specific batch: (a) B01 (including B01b); (b) B02; (c) B03; and (d) B04.

### S4. Supporting data about the economic analysis of the crutches

Detailed costs are available on OSF. The methodology followed for the analysis is explained in the main text (Section "Economic analysis"). The nominal weights and 3D printing times of the custom 3D-printed parts are listed in Table S5, together with the total weights and times, divided per batch (material and 3D printing scale).

Table S5. Nominal weight and 3D printing times of the 3D printed crutch parts produced with different materials, i.e., virgin and recycled PETG and virgin TPU, and 3D printer formats, i.e., small- and large-format, including total weights and times for each material and 3D printer.

| Part | Q.ty | Material | Small-scale FFF (B01, B02) | | Large-scale FFF* (B03, B04) | |
|---|---|---|---|---|---|---|
| | | | Weight (g) | Time (min) | Weight (g) | Time (min) |
| C01. Cuff body | 1 | Virgin PETG (B01, B03) | 114 | 207 | 114 | 209 |
| | | Recycled PETG (B02, B04) | 109 | | 109 | |
| H01. Handle body | 1 | Virgin PETG (B01, B03) | 220 | 277 | 220 | 276 |
| | | Recycled PETG (B02, B04) | 210 | | 210 | |
| B03. Food body | 1 | Virgin PETG (B01, B03) | 36 | 70 | 36 | 67 |
| | | Recycled PETG (B02, B04) | 35 | | 35 | |
| B05. Foot base | 1 | Virgin PETG (B01, B03) | 25 | 40 | 25 | 42 |
| | | Recycled PETG (B02, B04) | 24 | | 24 | |
| Total (PETG) | | Virgin PETG (B01, B03) | 396 | 594 | 395 | 594 |
| | | Recycled PETG (B02, B04) | 378 | | 377 | |
| C02. Cuff grip | 1 | TPU 90 Shore A | 26 | 278 | // | // |
| H02. Handle grip | 1 | TPU 90 Shore A | 27 | 299 | // | // |
| B02. Washer | 1 | TPU 90 Shore A | 1 | 12 | // | // |
| B04. Living joint | 1 | TPU 90 Shore A | 11 | 122 | // | // |
| B06. Foot base grip | 1 | TPU 90 Shore A | 11 | 119 | // | // |
| Total (TPU) | | TPU 90 Shore A | 76 | 830 | 76 | 830 |
| Total (3D print) | | vPETG + TPU (B01, B03) | 472 | 1424 | 472 | 1424 |
| | | rPETG + TPU (B02, B04) | 454 | | 454 | |

**= Values calculated on a single component, starting from the gcode for multiple 3D printing.*

Table S6 shows the cost analysis of the 3D printed parts made in PETG and TPU, divided by batch, whereas Table S7 includes the overall costs of the assembly for each batch of crutches.

Table S6. Costs of the different crutch samples according to the different batches, showing the unit cost, total cost, and percentage cost for each batch and part, together with the total costs for a single crutch and a set of two. Prices are in USD.

| Part | Q.ty | Element | B01 | B02 | B03 | B04 |
|---|---|---|---|---|---|---|
| C01. Cuff body | 1 | Material (PETG) | 1.56 | 2.12 | 1.56 | 2.11 |
| | | Energy | 0.02 | 0.02 | 0.50 | 0.50 |
| | | Total | 1.58 | 2.14 | 2.06 | 2.61 |
| C02. Cuff grip | 1 | Material (TPU) | 1.35 | 1.35 | 1.35 | 1.35 |
| | | Energy | 0.03 | 0.03 | 0.03 | 0.03 |
| | | Total | 1.38 | 1.38 | 1.38 | 1.38 |
| H01. Handle body | 1 | Material (PETG) | 3.01 | 4.08 | 3.01 | 4.08 |
| | | Energy | 0.03 | 0.03 | 0.67 | 0.66 |
| | | Total | 3.04 | 4.11 | 3.68 | 4.74 |
| H02. Handle grip | 1 | Material (TPU) | 1.43 | 1.43 | 1.43 | 1.43 |
| | | Energy | 0.04 | 0.04 | 0.04 | 0.04 |
| | | Total | 1.47 | 1.47 | 1.47 | 1.47 |
| B02. Washer | 1 | Material (TPU) | 0.06 | 0.06 | 0.06 | 0.06 |
| | | Energy | >0.01 | >0.01 | >0.01 | >0.01 |
| | | Total | 0.06 | 0.06 | 0.06 | 0.06 |
| B03. Foot body | 1 | Material (PETG) | 0.50 | 0.67 | 0.50 | 0.67 |
| | | Energy | >0.01 | 0.01 | 0.16 | 0.17 |
| | | Total | 0.50 | 0.68 | 0.66 | 0.84 |
| B04. Living joint | 1 | Material (TPU) | 0.59 | 0.59 | 0.59 | 0.59 |
| | | Energy | 0.02 | 0.02 | 0.02 | 0.02 |
| | | Total | 0.61 | 0.61 | 0.61 | 0.61 |
| B05. Foot base | 1 | Material (PETG) | 0.35 | 0.47 | 0.35 | 0.47 |
| | | Energy | >0.01 | 0.01 | 0.10 | 0.10 |
| | | Total | 0.35 | 0.48 | 0.45 | 0.57 |
| B06. Foot base grip | 1 | Material (TPU) | 0.58 | 0.58 | 0.58 | 0.58 |
| | | Energy | 0.01 | 0.01 | 0.01 | 0.01 |
| | | Total | 0.59 | 0.59 | 0.59 | 0.59 |
| **Total** | | **PETG** | **5.48** | **7.42** | **6.84** | **8.77** |
| | | **TPU** | **4.11** | **4.11** | **4.11** | **4.11** |
| | | **Total 3D print** | **9.59** | **11.53** | **10.95** | **12.88** |

Table S7. Costs (in USD) of the different crutch samples according to the different batches, showing the unit cost, total cost, and percentage cost for each batch and part.

| Part | Q.ty | Cost | | | | | | | |
|---|---|---|---|---|---|---|---|---|---|
| | | B01 | | B02 | | B03 | | B04 | |
| | | Per unit | Total | Per unit | Total | Unit | Total | Unit | Total |
| C01. Cuff body | 1 | 1.58 | 1.58 | 2.14 | 2.14 | 2.06 | 2.06 | 2.61 | 2.61 |
| C02. Cuff grip | 1 | 1.38 | 1.38 | 1.38 | 1.38 | 1.38 | 1.38 | 1.38 | 1.38 |
| C03. Upper dowel | 1 | 1.20 | 1.20 | 1.20 | 1.20 | 1.20 | 1.20 | 1.20 | 1.20 |
| H01. Handle body | 1 | 3.04 | 3.04 | 4.11 | 4.11 | 3.68 | 3.68 | 4.74 | 4.74 |
| H02. Handle grip | 1 | 1.47 | 1.47 | 1.47 | 1.47 | 1.47 | 1.47 | 1.47 | 1.47 |
| B01. Lower dowel | 1 | 4.79 | 4.79 | 4.79 | 4.79 | 4.79 | 4.79 | 4.79 | 4.79 |
| B02. Washer | 1 | 0.06 | 0.06 | 0.06 | 0.06 | 0.06 | 0.06 | 0.06 | 0.06 |
| B03. Foot body | 1 | 0.50 | 0.50 | 0.68 | 0.68 | 0.66 | 0.66 | 0.84 | 0.84 |
| B04. Living joint | 1 | 0.61 | 0.61 | 0.61 | 0.61 | 0.61 | 0.61 | 0.61 | 0.61 |
| B05. Foot base | 1 | 0.35 | 0.35 | 0.48 | 0.48 | 0.45 | 0.45 | 0.57 | 0.57 |
| B06. Foot base grip | 1 | 0.59 | 0.59 | 0.59 | 0.59 | 0.59 | 0.59 | 0.59 | 0.59 |
| F01. Wood screw | 13 | 0.07 | 0.92 | 0.07 | 0.92 | 0.07 | 0.92 | 0.07 | 0.92 |
